# Hitchhiker bias can distort inference of pathogen severity and pathogen–pathogen interactions from symptom-based surveillance: A modeling study

**DOI:** 10.64898/2026.03.13.26348325

**Authors:** Kiran Kumari, Sarah C. Kramer, Matthieu Domenech de Cellès

**Affiliations:** Max Planck Institute for Infection Biology, Infectious Disease Epidemiology group, Berlin, Germany

**Keywords:** Infectious disease surveillance, transmission models, observation models, statistical models, selection bias, co-infection, multiplex PCR

## Abstract

Symptom-based infectious disease surveillance, which infers pathogen circulation by testing individuals who seek care for clinical symptoms, is widely used in public health but can distort our understanding of pathogen dynamics. Detecting a pathogen in a hospitalized patient does not establish that it caused the hospitalization. When a mildly pathogenic virus is detected in patients hospitalized because of a co-infecting, more pathogenic virus, the hospitalization can be wrongly attributed to the mild virus, a mechanism we term hitchhiker bias. Building on the aetiology-attribution literature, we couple this misattribution to a two-pathogen SEIR model with explicit co-infection and multiplex observation, and ask how an unmodeled incidental-detection pathway is absorbed into the parameters estimated by transmission models. We show that the relative inflation of observed hospitalizations is governed by the ratio of the two viruses’ hospitalization probabilities and by the fraction of mild-virus infections that are co-infections. In simulation–estimation experiments, ignoring the hitchhiker pathway overestimated the hospitalization probability of the mild virus about 3-fold under partial and 7-fold under complete epidemic overlap, with credible intervals that excluded the true value. In the absence of biological interaction, it also overestimated the mild virus’s reproduction number and generated spurious inhibition under partial overlap, and generated spurious facilitation under complete overlap. When the viruses truly interacted, the bias in the estimated interaction depended on epidemic overlap: a true inhibition appeared stronger under partial overlap but was reversed into weak facilitation under complete overlap, and a true facilitation appeared stronger. Explicitly modeling the hitchhiker pathway recovered hospitalization probabilities and interaction strengths when the observation model was correctly specified. These results highlight the importance of separating pathogen-attributable hospitalization from incidental detection when drawing epidemiological inferences from symptom-based surveillance.

**Author summary:** When people are sick enough to go to hospital, doctors can now test them for many viruses at once. It is easy to assume that any virus found in a patient helped cause the hospital stay. But that is not always true. A patient can carry a mild virus that causes few symptoms and also a more serious virus that is the real reason they were admitted. The mild virus is then found only because the patient happened to be tested for the serious one. We call this “hitchhiker bias”. Using computer simulations of two viruses spreading through a population, we found that this can make a mild virus look more serious than it really is. It can also make the two viruses look as if they help each other spread when they do not. The effect is strongest when both viruses are spreading at the same time. In our simulations, a method that took this into account gave far more accurate answers.

## Introduction

Infectious diseases remain a major global health challenge, causing millions of deaths and substantial disability each year [1–3]. Symptom-based surveillance systems, which test patients who seek care rather than the population at large, are essential for monitoring pathogen circulation, co-infection patterns and pathogen interactions. Multiplex molecular panels now allow many pathogens to be tested in a single clinical encounter, and the resulting data on pathogens detected in symptomatic patients are growing rapidly [4,5]. Inference from these data rests on an assumption that is seldom stated: that a pathogen detected in a patient contributed to the clinical event that brought the patient into surveillance.

Aetiology research has long shown that this assumption cannot be taken for granted, because pathogens are routinely detected in people without disease. The Global Enteric Multicenter Study (GEMS) used matched asymptomatic controls to estimate the fraction of pediatric diarrhea attributable to each pathogen [6]. In the CDC Etiology of Pneumonia in the Community (EPIC) study, multiple pathogens were detected in about a quarter of hospitalized children with pneumonia [7], and a comparison with asymptomatic controls showed that the significance of a viral detection depends on the virus and on the age of the patient [8]. These findings motivated statistical methods that integrate case and control data while accounting for imperfect measurements, including partially latent class models and Bayesian integrated analyses [9]. Their conclusions can depend on the diagnostic approach: quantitative PCR reanalysis of the diarrhea study raised the pathogen-attributable fraction of cases from 51.5% to 89.3% [10]. In the multicountry Pneumonia Etiology Research for Child Health (PERCH) study, almost all cases (98.9%) and community controls (98.0%) had at least one pathogen detected by PCR in nasopharyngeal–oropharyngeal specimens, so aetiology had to be estimated by integrating case–control data from multiple specimens [11].

The same problem arises when a single pathogen is detected among admitted patients. In a chart review of SARS-CoV-2-positive admissions in four US health systems, about a quarter were incidental, that is, patients admitted with rather than for COVID-19 [12], and national hospital analyses accordingly distinguished admissions primarily for COVID-19 from the rest [13].

As multiplex panels spread, co-detections have become routine observations, but they remain difficult to interpret. Meta-analytic evidence on whether viral co-infection worsens clinical disease is inconclusive [14], and apparent changes in pathogen-specific burden can follow changes in testing practice [15], as when the introduction of multiplex panels changed the ascertainment of pertussis-attributable deaths [16]. Surveillance samples are also subject to selection: restricting analyses to patients who are ill enough to seek care or be admitted (collider bias) [17–19] can induce spurious associations between pathogens that are independent in the community, a concern raised specifically for co-detection studies [20,21]. Healthcare-seeking behavior and imperfect test performance [22–26] and shared seasonality [27–29] further complicate interpretation, and co-detection prevalence is a poor guide to interaction even in the absence of selection [30].

For mechanistic transmission models, the observation process that links infection to surveillance data matters as much as the transmission process: biases in surveillance data can propagate to estimates of the reproduction number [31]. Eales et al. [32] developed a mathematical description of case-based surveillance and showed that common indicators, such as case counts and the test-positive proportion, can be heavily biased by co-circulating pathogens with similar symptom profiles, in part because people infected with the target pathogen may be tested because of symptoms generated by another process. Their analysis centered on surveillance indicators. How such misattribution propagates into the parameters of mechanistic models fitted to the data, notably severity and pathogen interaction, has not been characterized.

Mechanistic models are the main tool for inferring pathogen interactions from surveillance data. Simulation studies show that, when the epidemiological and observation processes are well specified, time series can carry enough information to identify interactions [33], but identifiability can fail toward the extremes of plausible parameter ranges [33,34]. Interactions [35] can alter incidence trends and the interpretation of surveillance [36], and co-infections by non-interacting pathogens need not be statistically independent [37]. Models of this kind have been fitted to multiplex clinical data [38] and to influenza and RSV surveillance data [39], whereas simpler non-mechanistic methods remain unreliable [39,40]. Some have been fitted directly to hospital-based multiplex counts [35]. These models treat each pathogen-specific detection as a hospitalization due to that pathogen. An incidental-detection pathway could produce a similar signature, because the second pathogen is detected in patients admitted because of the first, yet whether and how such a pathway is absorbed into estimated parameters has not been examined.

We call this mechanism hitchhiker bias. It occurs when a mildly pathogenic or asymptomatic pathogen is detected in a co-infected patient because another, more pathogenic infection caused the hospitalization that triggered testing, and the detection is then attributed to the detected pathogen. Selection on admission (a collider) is followed by misattribution of the admission to the detected pathogen (a misclassification); hitchhiker bias refers to the second step. When infections are independent, the probability that the milder virus is detected among patients admitted because of the severe virus equals its community prevalence, so it is the attribution of that detection, not the detection itself, that is biased. Collider bias therefore distorts associations between pathogens through sample selection, whereas hitchhiker bias distorts the mapping from pathogen detection to pathogen-attributable disease burden.

Here we ask how hitchhiker bias distorts inference from hospital-based multiplex surveillance and whether an explicit observation model can correct it. We couple a two-pathogen SEIR model with explicit co-infection to a multiplex observation process that separates pathogen-attributable hospitalizations from pathogen detections among hospitalized patients. We first derive how the relative inflation of observed hospitalizations depends on the relative severity of the two viruses and on the co-infected fraction of the milder virus’s prevalence. We then simulate epidemics from minimal to complete temporal overlap to characterize the amplitude and timing of the bias. Next, we fit transmission models to synthetic hospitalization data, with and without true biological interaction, to determine whether the unmodeled pathway is absorbed into reproduction numbers, pathogenicity, or interaction strength. Finally, we test whether an adjusted observation model recovers the underlying parameters. Our contribution is threefold: (i) we couple pathogen attribution to a two-pathogen transmission model with explicit co-infection; (ii) we show which mechanistic parameters absorb the misspecification and how this depends on epidemic overlap and relative pathogenicity; and (iii) we evaluate, in simulation, an adjusted observation model that represents hitchhiking detections.

## Results

To characterize the hitchhiker mechanism in its simplest form, we first considered two co-circulating viruses with no biological interaction (θ = 1, i.e., infection with one virus does not change susceptibility to the other) that differed only in their probability of causing hospitalization, denoted by ρ. Virus 1 was specified as the more pathogenic virus (ρ_1_ = 0.05) and virus 2 as the less pathogenic virus (ρ_2_ = 0.0005), corresponding to a severity ratio κ = ρ_1_/ρ_2_ = 100. All other epidemiological parameters were identical for the two viruses (Table 1). We varied the relative timing of the two epidemics by changing the initial number of individuals exposed to virus 1 while keeping that of virus 2 fixed, which generated scenarios ranging from temporally separated to fully synchronized epidemics (Fig. 3a). For each scenario, we compared two quantities for each virus: the true hospitalizations, defined as hospitalizations attributable to that virus (H_i_), which are the epidemiological target of inference; and the observed hospitalizations, defined as virus-positive hospitalizations recorded by multiplex surveillance (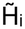), which are the quantities actually observed (Fig 3b). Because the simulations were deterministic and free of observation error, any discrepancy between H_i_ and 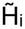 arose solely from the incidental detection of one virus in patients hospitalized because of the other. We first describe this discrepancy for three representative scenarios (minimal, partial, and complete overlap) and then quantify how it depends on epidemic overlap and relative severity.

**Table 1.** List of model parameters. The subscript *i* takes the value of 1 for virus 1 and 2 for virus 2.

| Parameters | Description | Values for virus 1 | Values for virus 2 |
|---|---|---|---|
| $\mathcal{R}_i$ | Basic reproduction number | 2.5 | 2.5 |
| ${\sigma_i}^{-1}$ | Average latent period | 4 days | 4 days |
| ${\gamma_i}^{-1}$ | Average infectious period | 5 days | 5 days |
| $R_{i,0}$ | Initial number of recovered individuals | 0 | 0 |
| $E_{i,0}$ | Initial number of exposed individuals | [$N/10^{12}$, $N/10^{2}$] | $N/10^{12}$ |
| $\rho_i$ | Hospitalization probability | 0.05 | 0.0005 |
| N | Population size | $10^6$ | $10^6$ |

### Co-infection with a more pathogenic virus inflates the observed burden of a mild virus

#### Minimal overlap

When the two epidemics were temporally separated, the epidemic of virus 1 had largely ended by the time virus 2 began to spread (Fig. 3a, left). Virus 1 peaked at day 40 and virus 2 at day 220, and their prevalence curves crossed only at very low levels, so co-infections were negligible. Almost every hospitalized patient who tested positive for virus 2 had therefore been hospitalized because of virus 2 itself, and the observed hospitalization curves were indistinguishable from the true curves for both viruses (Fig. 3b, left). In this setting, symptom-based surveillance gave an accurate picture of the low burden of virus 2, and hitchhiker bias was absent.

#### Partial overlap

When the virus-2 epidemic was initiated before the virus-1 epidemic had subsided, the simulations showed a clear signature of hitchhiker bias (Fig. 3b, middle). During co-circulation, a fraction of individuals infected with virus 2 were also infected with virus 1. In the model, a subset of these co-infected individuals was hospitalized because of virus 1, and multiplex testing then detected virus 2 incidentally. Because the standard observation model attributes every detection to the detected virus, these individuals were also recorded as virus-2 hospitalizations, even though virus 2 did not contribute to their hospitalization. This hitchhiker component (shaded area in Fig. 3b) inflated the observed virus-2 hospitalization curve. We quantified this amplitude distortion as the relative bias in peak hospitalizations, defined as [max(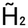) − max(H_2_)]/max(H_2_), irrespective of when each maximum occurred. A relative bias of 0 indicates no distortion, and a value of 1 indicates that the observed peak is twice the true peak. In this scenario, the observed peak reached ≈3.5 hospitalizations per 10^5^ population per day, compared with a true peak of ≈1.3, giving a relative bias of ≈1.7. Because virus 1 peaked earlier, the hitchhiker component was concentrated in the ascending phase of the virus-2 epidemic and advanced the observed peak. We quantified this temporal distortion as the difference between the times at which 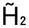 and H_2_ reached their maxima. The observed peak occurred ≈11 days before the true peak. Partial overlap therefore distorted both the magnitude and the timing of the observed virus-2 epidemic. We characterize both distortions across the full range of epidemic overlap in the next section.

#### Complete overlap

When the two epidemics were fully synchronized (Fig. 3a, right), the prevalence of co-infection was highest and the hitchhiker component made up most of the observed virus-2 hospitalizations (Fig. 3b, right). Because both viruses circulated at high prevalence over the same period, a large fraction of individuals infected with virus 2 were also infected with virus 1. Many of them were hospitalized because of virus 1 and then recorded as virus-2 hospitalizations following multiplex testing. The observed peak reached ≈17 hospitalizations per 10^5^ population per day, compared with a true peak of ≈1.3, giving a relative bias of ≈12.6. This was the largest amplitude distortion among the three scenarios, even though the intrinsic pathogenicity of virus 2 was the same in all of them. Unlike under partial overlap, the observed and true peaks occurred on the same day (peak shift ≈0 days). Because the hitchhiker component scales with the prevalence of co-infection, it rose and fell in step with the virus-2 epidemic. It therefore increased the amplitude of the observed curve and made it more sharply peaked, but did not shift its timing. Complete overlap thus produced the largest distortion in magnitude but none in timing, whereas partial overlap distorted both.

In all three scenarios, the observed and true hospitalizations of virus 1 were nearly identical (Fig. 3b). In principle, virus 1 can also hitchhike on virus 2: a patient hospitalized because of virus 2 may test positive for virus 1, and that detection would be recorded as a virus-1 hospitalization. This pathway is included in the model as the term (1 − ρ_1_)ρ_2_γ_2_NP_12_ in 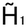 (Fig. 1). However, because virus 2 rarely causes hospitalization (ρ_2_ was 100-fold lower than ρ_1_), few such patients exist, and the contribution of this pathway was negligible. Under the conditions simulated here, hitchhiker bias was therefore strongly asymmetric. It distorted the observed dynamics of the less pathogenic virus and left those of the more pathogenic virus essentially unaffected. These three scenarios were chosen to illustrate the mechanism rather than to represent specific pathogen pairs or realistic epidemic timings. They show that, in this theoretical setting, the extent of hitchhiker bias depends on both the degree of epidemic overlap and the relative pathogenicity of the two viruses. We quantify these dependencies systematically in the next section.

**Figure 1.**
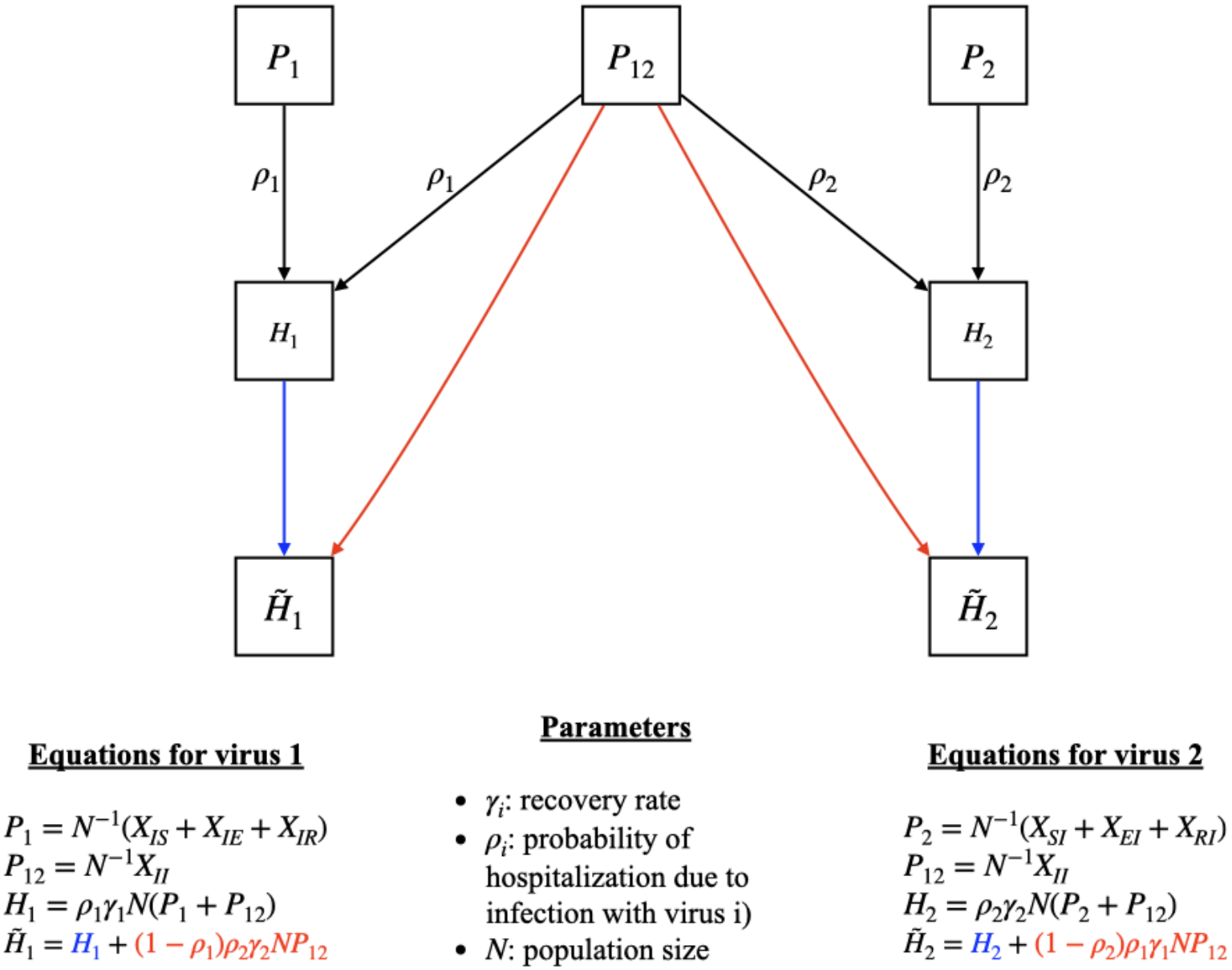
Diagram illustrating hitchhiker bias in symptom-based surveillance: The schematic shows how true hospitalizations for each virus (*H*_1_, *H*_2_), prevalence of co-infections (*P*_12_), and hospitalization probabilities (*ρ*_1_, *ρ*_*2*_)combine to produce the observed hospitalizations 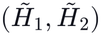. The blue arrows represent contributions from single infections of each virus, whereas the red arrows represent additional hospitalizations arising from co-infected individuals, highlighting how virus 2 can appear inflated in the observed data when it “hitchhikes” on hospitalizations driven primarily by virus 1.

### Hitchhiker bias depends on epidemic overlap and relative pathogenicity

To quantify hitchhiker bias systematically, we examined how the relationship between true (H_2_) and observed (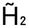) virus-2 hospitalizations changed as the two epidemics overlapped to different degrees. To describe this overlap with a single number, we calculated the Pearson correlation coefficient (r) between the prevalence of infection with virus 1, I_1_(t), and with virus 2, I_2_(t), over the simulation period (see Model and Methods). We refer to this quantity as epidemic overlap. A value of r = 1 means that the two prevalence curves rise and fall together; that is, the epidemics are fully synchronized. Lower values mean that the epidemics are increasingly separated in time. Across our simulations, epidemic overlap ranged from r =−0.11 (95% confidence interval [CI]: −0.20 to −0.02) for the most temporally separated epidemics considered (minimal overlap) to r = 1 for fully synchronized epidemics (complete overlap). The lowest value did not approach −1, as might be expected for non-overlapping epidemics. A correlation of −1 would require one curve to rise exactly as the other falls. Two separated epidemics, by contrast, are both close to zero for most of the simulation period, before and after their respective peaks. These shared periods of low prevalence keep their correlation near zero, even when their peaks are far apart. The value r = −0.11 therefore represents the lowest overlap achieved in our simulations rather than a theoretical minimum. Values between −0.11 and 1 correspond to progressively greater temporal alignment of the two epidemics.

#### Amplitude Distortion

We first asked how much hitchhiker detections inflate the observed number of virus-2 hospitalizations. As described above, we measured this inflation as the relative bias in peak hospitalizations, [max(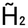) − max(H_2_)]/max(H_2_), which expresses the excess of the observed peak over the true peak as a proportion of the true peak. For a severity ratio of κ = 100, the relative bias increased steadily as the two epidemics became more synchronized (Fig. 4a). Under minimal overlap (point A), it was close to zero, meaning that the observed and true peaks were almost identical. Under partial overlap (point B), it reached ≈1.7, so the observed peak was nearly three times the true peak. Under complete overlap (point C), it reached ≈12.6, so the observed peak was more than an order of magnitude higher than the true peak.

This pattern follows directly from the structure of the observation model (Fig. 1). At any time point, the relative inflation of observed over true virus-2 hospitalizations is given by

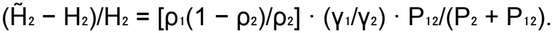

Because the two viruses have equal recovery rates in our simulations (γ_1_ = γ_2_), this expression simplifies to

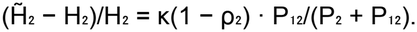

The bias is therefore the product of two terms, each with a simple interpretation. The first, κ(1− ρ_2_), is a severity term. It describes how much more likely virus 1 is than virus 2 to cause hospitalization, and is approximately equal to κ when virus 2 is mild. The second, P_12_/(P_2_ + P_12_), is an overlap term. It is the proportion of individuals infected with virus 2 who are also infected with virus 1 at that time. This proportion is close to zero when the epidemics are separated in time, and it increases as they become synchronized. This decomposition also explains why the bias was largest for a mild virus. Because virus 2 rarely causes hospitalization on its own, its true hospitalizations (H_2_) are small. Even a modest number of incidental detections in patients hospitalized because of virus 1 is therefore large relative to H_2_, and dominates its observed signal.

#### Temporal Bias in Peak Detection

Hitchhiker detections also changed the apparent timing of the virus-2 epidemic. We measured this effect as the peak shift, defined as the difference between the day on which observed virus-2 hospitalizations (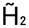) reached their maximum and the day on which true virus-2 hospitalizations (H_2_) reached theirs. Negative values indicate that the observed peak occurs earlier than the true peak. For κ = 100, the peak shift did not change monotonically with epidemic overlap (Fig. 4b). Under minimal overlap, the observed and true peaks coincided. As overlap increased, the observed peak moved progressively earlier, reaching a maximum advance of ≈11 days at partial overlap (epidemic overlap, r ≈ 0.15–0.25). With further increases in overlap, the shift became smaller again and disappeared when the epidemics were fully synchronized (r = 1).

This non-monotonic pattern can be understood by viewing the observed curve as the sum of two components: the true virus-2 hospitalizations and the hitchhiker component arising from co-infected patients hospitalized because of virus 1. The hitchhiker component depends on co-infection prevalence, which is highest when both viruses circulate at high levels. When virus 1 peaks before virus 2, the prevalence of co-infection, and therefore the hitchhiker component, peaks earlier than the true virus-2 curve. Adding an earlier-peaking component to the true curve shifts the maximum of the observed curve to an earlier time. The size of this shift depends on two opposing factors. The first is how large the hitchhiker component is relative to the true curve. The second is how far apart the peaks of the two components are. Under minimal overlap, the peaks are far apart, but the hitchhiker component is negligible, so the observed peak is not displaced. Under complete overlap, the hitchhiker component is large but peaks at the same time as the true curve, so it raises the observed peak without moving it. Only at intermediate overlap is the hitchhiker component both substantial and time-shifted relative to the true curve, and this is where the largest shifts occurred. The stepwise appearance of the curve in Fig. 4b reflects the daily resolution at which peak times were recorded.

Hitchhiker bias can therefore distort not only how large an epidemic of a mild virus appears in surveillance data, but also when it appears to peak. Amplitude and timing distortions were maximal under different conditions: amplitude bias was greatest when the epidemics were fully synchronized, whereas timing bias was greatest when they overlapped partially. An earlier apparent peak may be interpreted as faster epidemic growth. As shown below, this has consequences for the estimation of transmission parameters when models are fitted to observed data.

#### Dependence on relative pathogenicity

The analyses above assumed a strong contrast in pathogenicity between the two viruses (κ = 100). In practice, co-circulating pathogens can differ in severity to varying degrees. We therefore examined how hitchhiker bias changes as this contrast is reduced. Keeping the hospitalization probability of virus 1 fixed (ρ_1_ = 0.05), we varied that of virus 2 from ρ_2_ = 0.0005 to ρ_2_ = 0.05, corresponding to severity ratios from κ = 100, where virus 2 is much milder than virus 1, to κ = 1, where both viruses are equally likely to cause hospitalization. For each combination of severity ratio and epidemic overlap, we calculated the relative bias in peak hospitalizations (Fig. 4c) and the peak shift (Fig. 4d) for virus 2. In both panels, the horizontal axis shows epidemic overlap, the vertical axis shows the severity ratio, and the color scale and contour lines show the size of the distortion. Each contour line connects combinations of overlap and severity ratio that produce the same level of distortion.

The relative bias increased with both epidemic overlap and severity ratio (Fig. 4c). It was smallest in the lower-left part of the panel, where the epidemics were separated in time, and the two viruses had similar pathogenicity. It was largest in the upper-right corner, where the epidemics were synchronized, and virus 2 was much milder than virus 1 (relative bias ≈12.6). As κ approached 1, the bias became small at every level of overlap. When the two viruses are equally pathogenic, virus 2 causes a substantial share of hospitalizations itself, so incidental detections make up only a small fraction of its observed hospitalizations. The shape of the contour lines follows from the decomposition derived above. Because the relative bias is approximately the product of a severity term (≈κ) and an overlap term (the co-infected fraction of virus-2 infections), lines of equal bias are approximately hyperbolic. A reduction in one factor can therefore be offset by an increase in the other. For example, a moderate difference in pathogenicity combined with fully synchronized epidemics can produce the same bias as a large difference in pathogenicity combined with only partial overlap. Neither epidemic timing nor relative severity alone is therefore sufficient to judge whether hitchhiker bias is likely to be important; both must be considered together.

The peak shift showed a different pattern (Fig. 4d). For every severity ratio, the shift was close to zero at both extremes of overlap and largest at low-to-intermediate overlap (r ≈ 0.1–0.4), reproducing the non-monotonic pattern described above for κ = 100. The magnitude of this shift depended on the severity ratio. When κ was close to 1, peak shifts were negligible, because the hitchhiker component was too small relative to the true curve to displace its maximum. As virus 2 became milder, the hitchhiker component grew relative to the true curve and pulled the observed peak increasingly earlier. The largest advances, of ≈10–12 days, occurred at high severity ratios (κ ≳ 70) and low-to-intermediate overlap. At high severity ratios, the shift increased only slowly with κ. Once the hitchhiker component dominates the observed curve, the observed peak approaches the co-infection prevalence peak. The maximum possible shift is therefore limited by the time between the peak of co-infection and the true virus-2 peak, which is set by epidemic overlap rather than by severity. As in Fig. 4b, the stepwise contours reflect the daily resolution of peak times.

Together, these results show that, in this theoretical setting, relative pathogenicity and epidemic overlap play distinct roles in hitchhiker bias. The low pathogenicity of virus 2 determines how strongly its observed dynamics are distorted. Epidemic overlap determines how the distortion appears: as an inflated peak when the epidemics are synchronized, as an earlier peak when they partially overlap, or as both. Because these two forms of distortion affect different features of the observed epidemic curve, they may also bias different parameters when transmission models are fitted to surveillance data. We examine this question next.

### Ignoring hitchhiker detections biases estimates of pathogenicity, transmissibility and interaction

We next asked whether the distortions described above carry over into the parameters estimated when transmission models are fitted to surveillance data. For each of the three overlap scenarios (minimal, partial and complete), we generated 100 synthetic datasets of observed daily hospitalizations for both viruses. Observation noise was added by drawing daily counts from a negative binomial distribution whose mean equaled the simulated observed hospitalizations 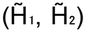, with dispersion parameter 0.04. We then fitted two models to each dataset by MCMC. In the first model, the hitchhiker effect was ignored (red in Fig. 5): every detection of a virus in a hospitalized patient was treated as a hospitalization caused by that virus. In the second model, the hitchhiker effect was modeled (teal in Fig. 5): the observation process explicitly included incidental detections in patients hospitalized because of the other virus. Both models estimated the same six parameters: the basic reproduction numbers (R_01_, R_02_), the hospitalization probabilities (ρ_1_, ρ_2_), the interaction parameter (θ) and the negative binomial dispersion. In every case, the data were generated without biological interaction (θ = 1). Fig. 5 shows, for each replicate, the posterior mean and 95% credible interval. Density curves summarize the distribution of posterior means across replicates.

#### Reproduction number of virus 1

Estimates of the basic reproduction number of the more pathogenic virus were largely unaffected by the hitchhiker effect (Fig. 5, first row). Under minimal and partial overlap, posterior means from both models were centered on the true value (R_01_ = 2.50), and the credible intervals of almost all replicates included it. The precision of these estimates increased with overlap: credible intervals spanned roughly 2.45–2.55 under minimal overlap and 2.49–2.51 under partial overlap. Under complete overlap, the model accounting for the hitchhiker effect still recovered the true value. The model ignoring it, by contrast, slightly but consistently underestimated R_01_ (posterior means ≈2.48), and most credible intervals excluded the true value. This deviation was small, less than 1% of the true value. However, the observed virus-1 data were themselves essentially unbiased (Fig. 3b), so it cannot reflect incidental detections of virus 1. It instead points to a compensatory adjustment linked to the bias in other parameters, which we discuss below.

#### Pathogenicity of virus 1

A similar pattern was observed for the hospitalization probability of virus 1 (ρ_1_; Fig. 5, second row). Under minimal and partial overlap, both models recovered the true value (ρ_1_ = 0.05). Under complete overlap, the model ignoring the hitchhiker effect overestimated ρ_1_ (posterior means ≈0.058, roughly 16% above the true value), whereas the model accounting for it remained centered on the true value.

#### Reproduction number of virus 2

The basic reproduction number of virus 2 was affected mainly under partial overlap (Fig. 5, third row). In this scenario, the model ignoring the hitchhiker effect overestimated R_02_ (≈2.62 vs. 2.50), whereas the model accounting for it recovered the true value. This bias is consistent with the earlier apparent peak of the observed virus-2 curve under partial overlap (Fig. 4b). A model that ignores the hitchhiker effect can reproduce an earlier, steeper rise in observed hospitalizations by increasing transmissibility. Under complete overlap, where the observed peak was inflated but not shifted, R_02_ was only marginally underestimated (≈2.49). Under minimal overlap, both models gave slightly low but uncertain estimates (≈2.43), with credible intervals that generally included the true value.

#### Pathogenicity of virus 2

The clearest consequence of ignoring the hitchhiker effect was on the estimated hospitalization probability of virus 2 (ρ_2_; Fig. 5, fourth row). Under minimal overlap, both models recovered the true value (ρ_2_ = 0.0005). As overlap increased, the model ignoring the hitchhiker effect increasingly overestimated ρ_2_. Posterior means clustered around ≈0.0015 under partial overlap and ≈0.0034 under complete overlap, approximately 3-fold and 7-fold the true value, respectively. These estimates were precise: their credible intervals were narrow and excluded the true value. When the hitchhiker effect was ignored, incidental detections were therefore attributed to the intrinsic pathogenicity of virus 2, and the model inferred, with high apparent confidence, that virus 2 was considerably more severe than it was. When the hitchhiker effect was modeled, estimates of ρ_2_ were close to the true value in all three scenarios.

#### Interaction

Ignoring the hitchhiker effect also produced spurious evidence of interaction between the two viruses, even though none was present in the simulations (Fig. 5, fifth row). The direction of this bias depended on epidemic overlap. Under complete overlap, the model ignoring the hitchhiker effect estimated θ ≈ 2.1, implying that infection with one virus roughly doubles susceptibility to the other. Under partial overlap, it estimated θ ≈ 0.7, implying a moderate inhibitory interaction. In both scenarios, the model accounting for the hitchhiker effect produced estimates close to the true value (θ = 1). Under minimal overlap, both models gave similar, highly uncertain estimates (posterior means ≈2.3; credible intervals spanning roughly 0.3–8). With almost no co-infections in this scenario, the data carry little information about interaction, and the estimates largely reflect the prior. Spurious interactions are particularly problematic because they could lead to incorrect conclusions about pathogen ecology. Depending on epidemic timing, an unmodeled hitchhiker effect can make two non-interacting viruses appear either to facilitate or to inhibit each other.

#### Propagation of bias across parameters

The biases in different parameters appeared to be linked. Under partial overlap, the overestimation *ℛ*_i1_ of was accompanied by an underestimation of θ. This may reflect the model reproducing the early rise of the observed virus-2 curve through higher transmissibility, while reducing the interaction to avoid overpredicting later co-infections. Under complete overlap, the excess virus-2 detections were absorbed mainly by ρ_2_ and θ. The spurious facilitation inferred in this scenario increases co-infection and therefore alters the simulated dynamics of virus 1, which the model offsets by slightly lowering *ℛ*_i1_ and raising ρ_1_. Bias arising from ignoring the hitchhiker effect for one pathogen can therefore propagate to estimates for the other, even when the data for that other pathogen are unbiased.

Together, these analyses show that, in our simulations, ignoring the hitchhiker effect biased several mechanistic parameters, and that which parameters were affected depended on epidemic overlap. Under partial overlap, where the main distortion was an earlier observed peak, the bias appeared in the transmissibility and pathogenicity of virus 2 and as spurious inhibition. Under complete overlap, where the main distortion was an inflated peak, the bias appeared mainly as overestimated pathogenicity of virus 2 and spurious facilitation, with smaller compensatory changes in the parameters of virus 1. In all scenarios, modeling the hitchhiker effect recovered parameter values close to the truth. Similar patterns were observed for other values of ρ_2_(0.005, 0.0125 and 0.025; S1–S3 Figs).

### Ignoring hitchhiker detections biases estimates of pathogenicity, transmissibility, and interaction in the presence of virus–virus interactions

The analyses above assumed that the two viruses did not interact. Respiratory viruses may, however, interact biologically. Innate immune responses triggered by one infection can reduce susceptibility to another, whereas tissue damage can increase it. We therefore examined whether a genuine interaction modifies hitchhiker bias, and whether it can still be estimated correctly from surveillance data. In our model, interaction is represented by the parameter θ, which scales susceptibility to one virus in individuals currently infected with the other (Fig. 2). Values of θ > 1 represent a positive interaction (facilitation), in which infection with one virus increases the risk of acquiring the other. Values of θ < 1 represent a negative interaction (inhibition), in which infection with one virus reduces that risk. θ = 1 represents no interaction. We considered a negative interaction (θ = 0.5) and a positive interaction (θ = 2) and compared them with the non-interacting case (θ = 1) under minimal, partial, and complete epidemic overlap. All other parameters, including the hospitalization probabilities (ρ_1_ = 0.05, ρ_2_ = 0.0005), were the same as in the previous sections.

**Figure 2:**
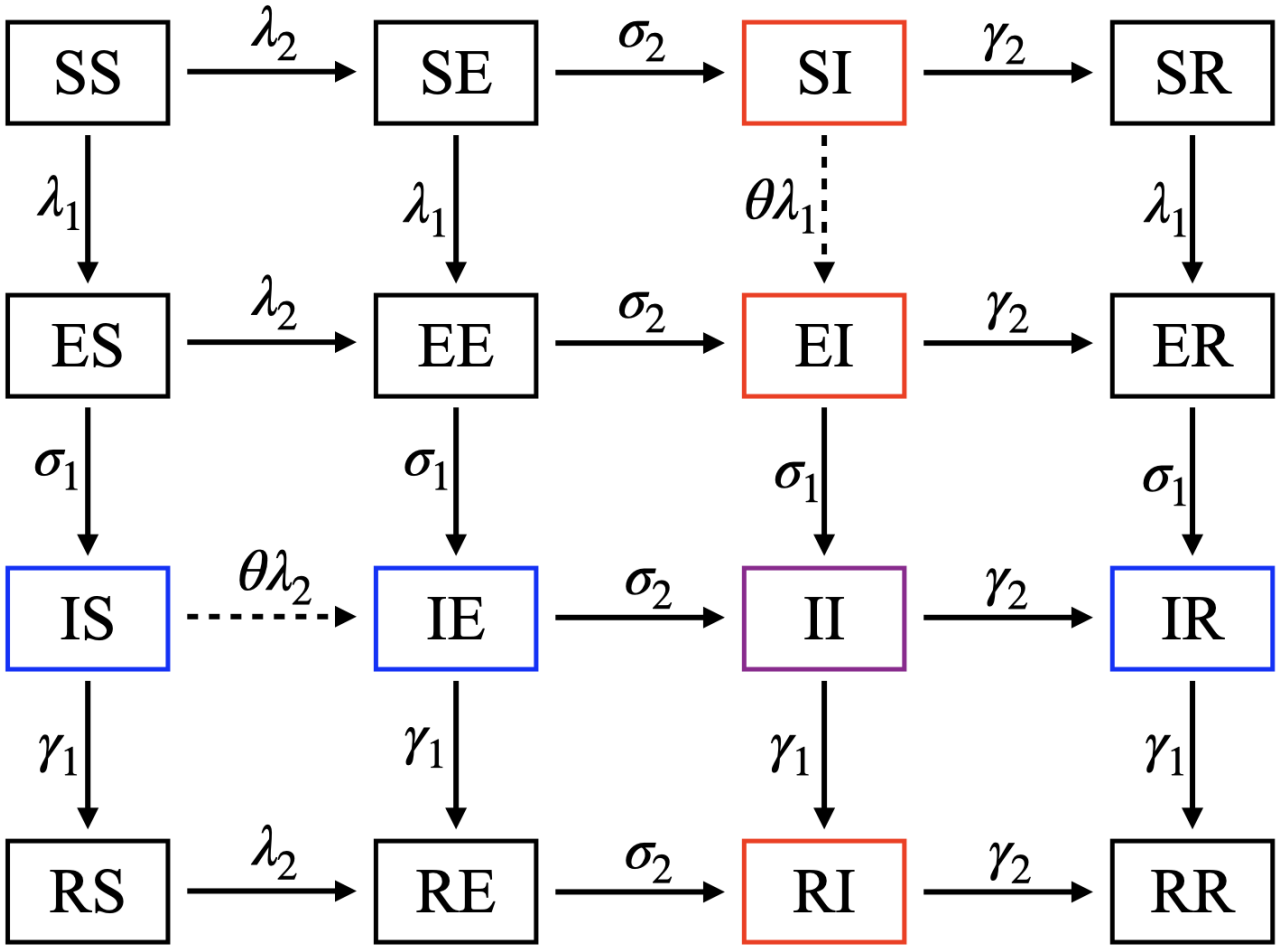
SEIR co-infection model for two viruses: Compartments represent the joint infection status of individuals, with the first and second letters indicating their status with respect to virus 1 (rows) and virus 2 (columns), respectively: susceptible (S), exposed (E), infectious (I), or recovered (R). Blue outlines identify compartments infectious with virus 1 only (IS, IE, and IR), red outlines identify compartments infectious with virus 2 only (SI, EI, and RI), and the purple outline identifies individuals infectious with both viruses (II). Black outlines indicate compartments in which individuals are not infectious with either virus. Arrows represent transitions between compartments; dotted arrows indicate acquisition of a second infection during an existing infectious episode, with susceptibility modified by the interaction parameter *θ*.

**Figure 3:**
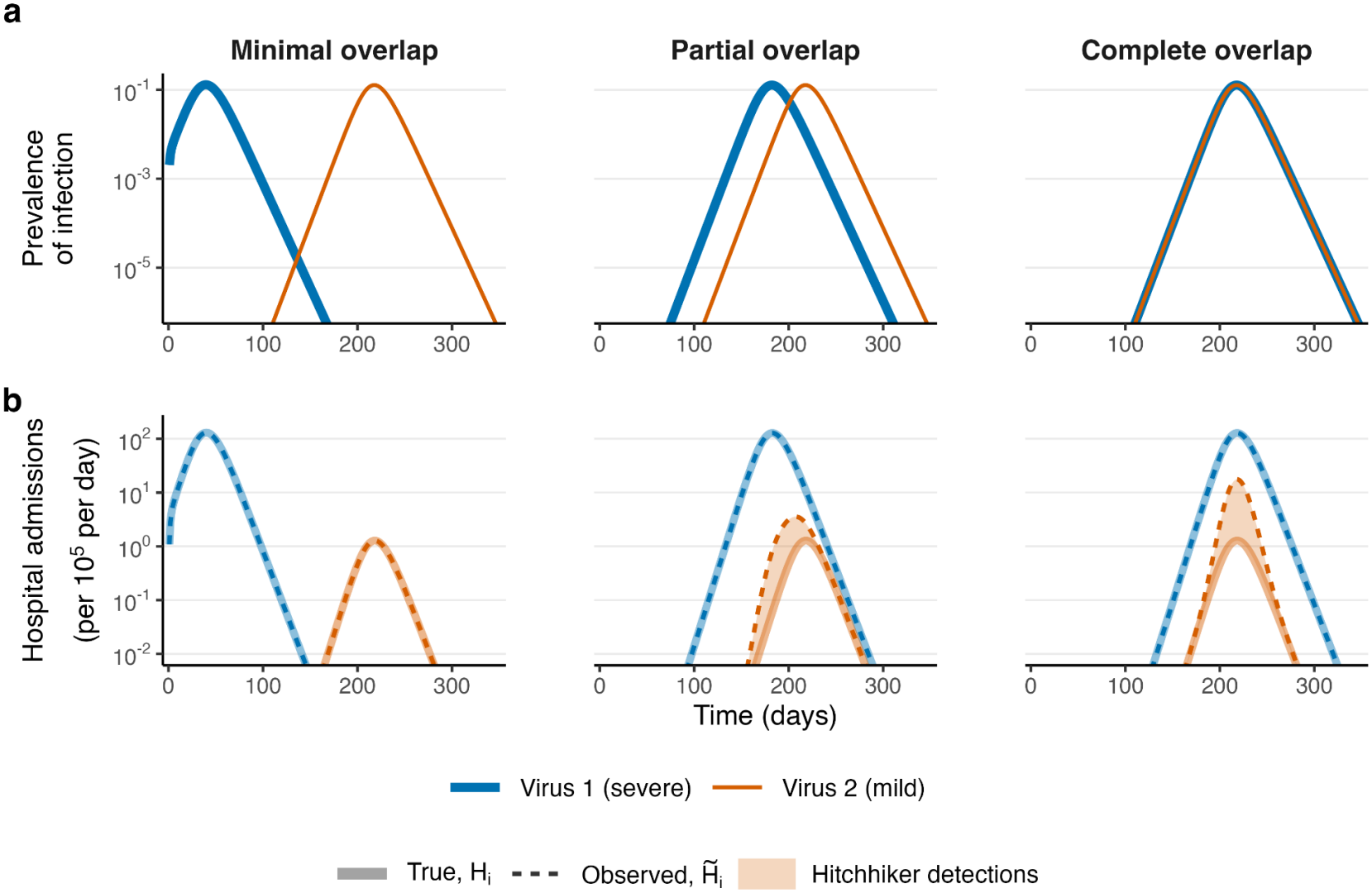
Hitchhiker bias under varying temporal overlap between two co-circulating viruses: (a) Prevalence of infection for the viruses 1 (blue) and virus 2 (orange) are shown across three distinct scenarios: a minimal overlapping infection curve (left panel), a partial overlap (middle panel), and a completely overlapped infection curve (right panel). (b) Comparison of the true and observed number of hospitalizations for both viruses under these different overlap conditions. The solid lines represent true hospitalizations (*H*_1_ *& H*_2_) and the dashed line represents the observed hospitalizations 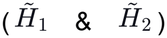. Observed hospitalizations for virus 2 (hitchhiker) increase as the infection curves of the two viruses overlap more.

#### Effect of interaction on observed hospitalizations

We first compared the true (H_2_) and observed 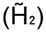 hospitalizations of virus 2 for each value of θ (Fig. 6a). Under minimal overlap, the observed and true curves coincided for all three values of θ. Because the two epidemics were separated in time, co-infections were rare regardless of interaction, leaving little opportunity for virus 2 to enter the surveillance data through hospitalizations caused by virus 1. As epidemic overlap increased, observed hospitalizations of virus 2 increasingly exceeded true hospitalizations, and the size of this excess depended on the interaction. Under partial overlap, the observed peak reached ≈2.2, ≈3.5 and ≈7 hospitalizations per 10^5^ population per day for θ = 0.5, 1 and 2, respectively, compared with a true peak of ≈1.3 in all cases. Under complete overlap, the corresponding observed peaks were ≈15, ≈17 and ≈25. These differences follow from the effect of interaction on the frequency of co-infection. A positive interaction increases the proportion of virus-2 infections that are co-infections with virus 1, and therefore the number of patients in whom virus 2 is detected incidentally after hospitalization because of virus 1. A negative interaction reduces this proportion and attenuates the bias. It did not, however, eliminate the bias, because co-infections still occurred whenever the epidemics overlapped. Interaction therefore modified the magnitude of hitchhiker bias but not its basic structure.

#### Estimation under interaction

We next asked whether the model parameters, including a true interaction, can be recovered when models are fitted to surveillance data. As in the non-interacting case, we generated 100 synthetic datasets with negative binomial observation noise for each combination of interaction (θ = 0.5 or 2) and epidemic overlap. To each dataset, we fitted two models, one in which the hitchhiker effect was ignored (red) and one in which it was modeled (teal). Replicates whose chains did not converge 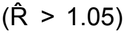 were excluded. Fig. 6b and 6c show the estimates of the two parameters most affected, the hospitalization probability of virus 2 (ρ_2_) and the interaction parameter (θ). Estimates of all five epidemiological parameters are shown in S4 Fig. (θ = 2) and S5 Fig. (θ = 0.5).

#### Pathogenicity of virus 2

As in the absence of interaction, ignoring the hitchhiker effect led to overestimation of ρ_2_ whenever the epidemics overlapped (Fig. 6b and 6c, upper rows). The magnitude of this overestimation depended on the interaction. Under a positive interaction, posterior means were ≈0.0023 under partial overlap and ≈0.0045 under complete overlap, approximately 5-fold and 9-fold the true value. Under a negative interaction, the corresponding values were ≈0.0012 and ≈0.0028, approximately 2.4-fold and 5.6-fold the true value. These values bracket those obtained without interaction (approximately 3-fold and 7-fold; Fig. 5). The bias in pathogenicity therefore increased with the strength of facilitation and decreased with the strength of inhibition, consistent with the corresponding changes in the number of incidental detections (Fig. 6a). Under minimal overlap, both models recovered ρ_2_ accurately. When the hitchhiker effect was modeled, ρ_2_ was recovered close to its true value in all scenarios.

#### Interaction

Ignoring the hitchhiker effect also biased the estimated interaction, and the direction of this bias depended on epidemic overlap (Fig. 6b and 6c, lower rows). Under complete overlap, θ was overestimated for both types of interaction. For a true positive interaction, the model ignoring the hitchhiker effect estimated θ ≈ 4.5, more than twice the true value. For a true negative interaction, it estimated θ ≈ 1.2, so that a truly inhibitory interaction appeared weakly facilitative. Under partial overlap, the pattern differed. For a true negative interaction, θ was underestimated (≈0.2), which exaggerated the inhibitory effect. For a true positive interaction, it was only slightly overestimated (≈2.3). When the hitchhiker effect was modeled, posterior estimates of θ were centered close to the true values under partial and complete overlap. Under partial overlap with a negative interaction, however, credible intervals were wide, indicating that the interaction was only weakly identifiable in that setting.

#### Other parameters

The remaining parameters showed the same pattern as in the absence of interaction (S4 and S5 Figs). Under minimal and partial overlap, both models recovered R_01_ and ρ_1_ close to their true values. Under complete overlap, the model ignoring the hitchhiker effect slightly underestimated R_01_ (≈2.48 for both θ = 0.5 and θ = 2) and overestimated ρ_1_. The overestimation of ρ_1_ increased with θ, from ≈0.056 for θ = 0.5 to ≈0.060 for θ = 2, compared with ≈0.058 without interaction. The reproduction number of virus 2 was overestimated under partial overlap (≈2.62 for θ = 0.5 and ≈2.60 for θ = 2), consistent with the earlier apparent peak of the observed virus-2 curve (Fig. 4b), and was close to the true value or slightly underestimated under complete overlap. The model accounting for the hitchhiker effect recovered all of these parameters close to their true values. The biases in transmissibility were therefore determined mainly by epidemic overlap and were nearly independent of the true interaction, and the bias in the pathogenicity of virus 1 varied only modestly with θ. By contrast, the biases in the pathogenicity of virus 2 and in the interaction parameter depended on both epidemic overlap and the true interaction, because the interaction changes the frequency of co-infection and hence the number of incidental detections of virus 2.

**Figure 4:**
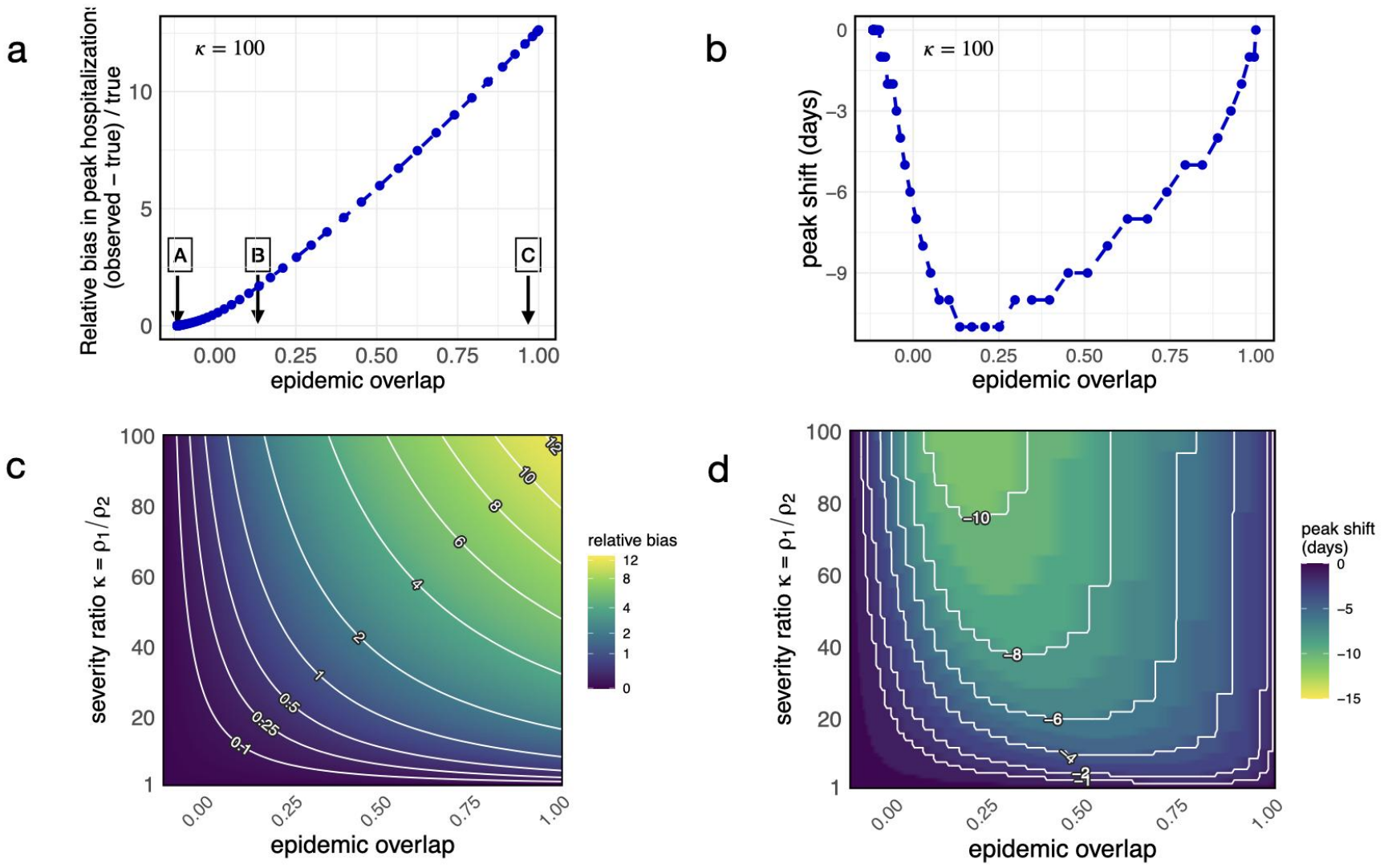
Hitchhiker bias depends on epidemic overlap and relative pathogenicity: (a) Relative bias in virus-2 hospitalizations, (observed − true)/true, as a function of epidemic overlap. Epidemic overlap is the Pearson correlation between the prevalences of virus 1 and virus 2, I_1_(t) and I_2_(t). Hospitalization probabilities were ρ_1_ = 0.05 and ρ_2_ = 0.0005 (severity ratio κ = ρ_1_/ρ_2_ = 100). Arrows A, B and C mark the minimal, partial and complete overlap scenarios shown in Fig 3. (b) Peak shift, defined as the day on which observed virus-2 hospitalizations 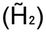 peak minus the day on which true virus-2 hospitalizations (H_2_) peak, for the same simulations as in (a). Negative values indicate that the observed peak occurs earlier than the true peak. (c, d) Relative bias (c) and peak shift in days (d) as functions of epidemic overlap and severity ratio. ρ_1_ was fixed at 0.05 and ρ_2_ was varied from 0.0005 to 0.05 (κ = 100 to 1). In both panels, contour lines connect combinations with equal values, as labeled.

**Figure 5:**
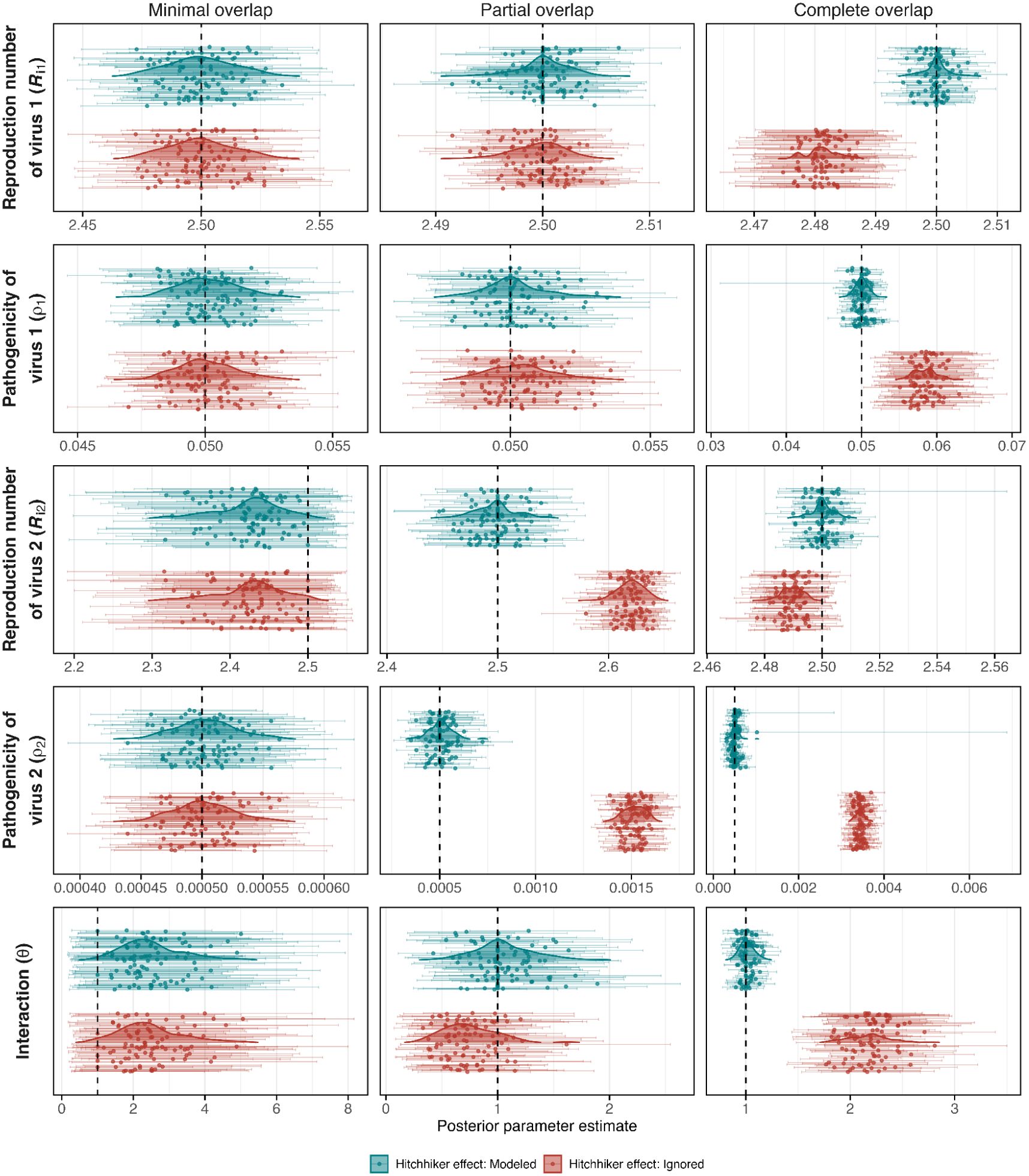
Ignoring hitchhiker detections biases estimates of pathogenicity, transmissibility, and interaction in the absence of virus–virus interactions: Posterior parameter estimates for simulated epidemics with minimal, partial, and complete epidemic overlap (columns). Rows show estimates of the basic reproduction number of virus 1, pathogenicity of virus 1, basic reproduction number of virus 2, pathogenicity of virus 2 and pathogen–pathogen interaction strength. Teal and red indicate estimates from models that account for or ignore the hitchhiker effect, respectively. Each point represents the posterior mean for one simulated replicate, and horizontal lines indicate the corresponding 95% credible intervals. Shaded density curves summarize the distribution of posterior means across simulated replicates. Vertical dashed lines indicate the true parameter values.

**Figure 6.**
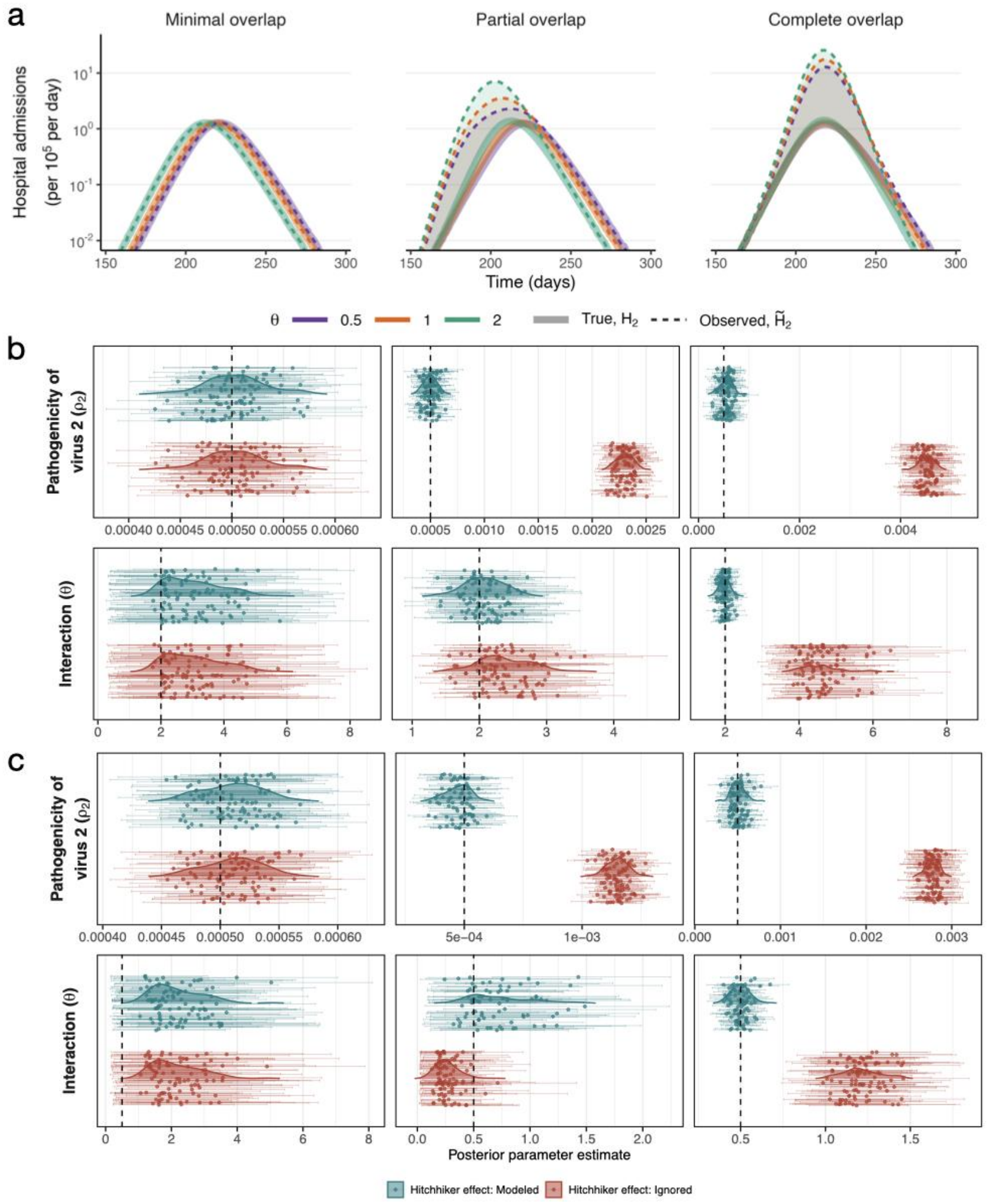
Ignoring hitchhiker detections biases estimates of pathogenicity, transmissibility, and interaction in the presence of virus–virus interactions: (a) True (H_2_, solid lines) and observed (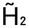, dashed lines) hospitalizations of virus 2 under minimal, partial and complete epidemic overlap, for a negative (θ = 0.5), absent (θ = 1), and positive (θ = 2) interaction. Shaded areas show the difference between observed and true hospitalizations, which is attributable to hitchhiker detections. (b, c) Posterior estimates obtained by fitting models in which the hitchhiker effect was modeled (teal) or ignored (red) to 100 simulated datasets per scenario, under (b) a positive interaction (θ = 2) and (c) a negative interaction (θ = 0.5). In each panel, the upper row shows the pathogenicity of virus 2 (ρ_2_) and the lower row shows the interaction parameter (θ). Each point is the posterior mean for one simulated dataset, and horizontal bars show the corresponding 95% credible interval. Shaded density curves summarize the distribution of posterior means across datasets. Vertical dashed lines indicate the true parameter values (ρ_2_ = 0.0005; θ = 2 in b and 0.5 in c). Estimates of all parameters are shown in S4 Fig (θ = 2) and S5 Fig (θ = 0.5).

These results show that hitchhiker bias distorts inference even when the pathogens genuinely interact. Whenever the epidemics overlapped, ignoring the hitchhiker effect overestimated the pathogenicity of the mild virus, and more so under facilitation than under inhibition. It also biased the estimated interaction. Under complete overlap, θ was overestimated roughly two-fold for every true value considered (0.5 → ≈1.2, 1 → ≈2.1, 2 → ≈4.5). A true facilitation therefore appeared stronger, an absence of interaction appeared as facilitation, and a true inhibition appeared as weak facilitation. Under partial overlap, a true inhibition was instead exaggerated. In addition, smaller biases in transmissibility and in the pathogenicity of the more pathogenic virus were reproduced consistently across interaction scenarios. If the observation process does not account for incidental detections, excess co-detections can thus be absorbed into several mechanistic parameters at once, and even the inferred direction of an interaction may be wrong. Explicitly modeling the hitchhiker effect separated biological interaction from incidental detection and recovered the underlying parameters in all scenarios in which the data were informative about interaction.

## Discussion

Multiplex testing of hospitalized patients is increasingly used to track the circulation, severity, and interactions of respiratory pathogens. Yet the data it generates record which pathogens are present, not which one caused the admission. By coupling a transmission model of two co-circulating viruses, in which individuals can carry both at once, with a multiplex-testing model, we showed that this gap can systematically misrepresent a mildly pathogenic virus that co-circulates with a more pathogenic one. Patients admitted because of the more pathogenic virus are tested, the milder virus is found incidentally, and the finding is counted as a hospitalization caused by the milder virus. In our simulations, this hitchhiker bias inflated the observed burden of the mild virus by more than an order of magnitude and advanced the apparent timing of its epidemic. When it was ignored during model fitting, it was absorbed into estimates of pathogenicity, transmissibility and interaction. A mild virus therefore appeared severe, and viruses that did not interact appeared to facilitate or inhibit one another. A model that represented incidental detection explicitly recovered the underlying parameters whenever the data contained information about them.

The size of the bias was governed by two quantities: the ratio of the hospitalization probabilities of the two viruses, and the proportion of infections with the milder virus that were co-infections with the more pathogenic one. Because the relative bias is approximately the product of these terms, a large difference in severity combined with modest epidemic overlap can be as distorting as a moderate difference combined with full synchrony (Fig. 4c). The bias should therefore be larger when the more pathogenic virus is more transmissible or more widespread. A less intuitive consequence is that low pathogenicity amplifies the bias rather than protecting against it. A virus that rarely causes admission on its own has a small true signal, which a modest number of incidental detections can overwhelm. Conversely, incidental detection of the more pathogenic virus in patients whose admission was caused by the milder one was negligible, so the bias was strongly asymmetric between the two pathogens.

Epidemic overlap affected the amplitude and the timing of the observed epidemic in different ways. Inflation of the observed peak increased steadily as the two epidemics became synchronized. The shift in peak timing, by contrast, was largest (up to about 11 days) when they overlapped only partly. This contrast reflects when incidental detections occur. When the more pathogenic virus peaks first, incidental detections accumulate in the rising phase of the mild-virus epidemic and pull its apparent peak earlier. When both viruses peak together, the same detections raise the observed curve without displacing it. The consequences of hitchhiker bias therefore depend on the timing of co-infections relative to each epidemic, not only on their number. Because epidemic timing is itself used to infer transmission and interaction, this temporal component matters for model-based inference as much as the inflation of burden.

When transmission models that ignored the hitchhiker effect were fitted to synthetic data, the hospitalization probability of the mild virus was the parameter most consistently affected. In the absence of interaction, it was overestimated roughly 3-fold under partial overlap and 7-fold under complete overlap. These estimates were precise, with credible intervals that excluded the true value. Other parameters absorbed the bias according to the way the observed curve was distorted. Under partial overlap, the earlier apparent peak was reproduced partly through a higher reproduction number of the mild virus. Under complete overlap, the inflated peak was reproduced mainly through higher pathogenicity and stronger interaction. Small compensatory shifts also appeared in the parameters of the more pathogenic virus, even though its own observations were essentially unbiased. In coupled transmission models, an error in observing one pathogen can therefore propagate to inference about the other.

Hitchhiker bias is related to collider bias but concerns a different inferential target. Collider bias arises when analyses are conditioned on healthcare seeking, testing, or hospitalization, because this conditioning can create associations between pathogens that are absent, or different, in the source population [17–21]. Hitchhiker bias concerns the interpretation of detections after this selection has taken place. A pathogen found in a hospitalized patient is treated as a cause of the admission, when the admission was in fact caused by another pathogen. The first bias therefore arises from who is included in the data, whereas the second arises from how their test results are attributed.

The two biases operate at different stages of the surveillance process and require different remedies. Collider bias is a property of analyses restricted to admitted patients. When two pathogens independently increase the risk of admission, they become associated among admitted patients even if they circulate independently in the community, and in the simplest case this association suggests inhibition rather than facilitation. Because collider bias requires both pathogens to influence admission, it is weakest when one rarely causes hospitalization, which is precisely when hitchhiker bias is strongest. Hitchhiker bias does not depend on any association being analyzed. It arises whenever pathogen-specific detections are used as a measure of admissions caused by that pathogen. In our simulations, it inflated estimated severity and, in most scenarios, generated apparent facilitation or inhibition depending on overlap. When both biases are present, they may therefore reinforce or partly offset each other, and correcting one does not correct the other. Collider bias is addressed by sampling from the source population or, where this is not possible, by analytical approaches such as weighting for the probability of selection [41]. Hitchhiker bias additionally requires information on the baseline probability of detecting each pathogen among patients admitted for other reasons, as provided by control groups in aetiological studies [6,11], or from an explicit observation model such as the one developed here.

From a methodological perspective, our model provides a proof of concept for correcting hitchhiker bias. When the observation process was correctly specified, representing incidental detection explicitly improved the recovery of pathogenicity, transmissibility, and interaction parameters in simulations. Applying this framework to surveillance data will require empirical validation and an assessment of its robustness when these conditions are not met. More broadly, three strategies could improve the reliability of inference from symptom-based surveillance. First, existing hospital-based data can continue to be used, with models such as ours applied to adjust explicitly for incidental detection. This adjustment is strengthened when routinely collected information helps to separate detection from attribution, such as the primary reason for admission [12] or quantitative measures of viral load [10]. Second, clinical surveillance can be complemented with random population-based sampling, which captures both symptomatic and asymptomatic infections and provides direct estimates of community prevalence. Third, hybrid designs can combine model-based correction with targeted community sampling or appropriate control groups [6,11], pairing the accuracy of population data with the feasibility of hospital-based surveillance.

Ignoring hitchhiker bias could also have practical consequences for public health. Mildly pathogenic or largely asymptomatic pathogens may be regarded as important causes of hospitalization, which could lead to misplaced concern, misallocation of clinical and diagnostic resources, and interventions targeted at the wrong pathogen. Spurious or distorted interaction estimates could, in turn, mislead expectations about the indirect effects of interventions, such as vaccination against one of two interacting viruses [39]. Accounting for incidental detection through model-based correction, expanded sampling, or a combination of both would give surveillance systems a more reliable basis for assessing pathogen burden and guiding policy.

Several limitations of our study should be noted. First, our model includes only two co-circulating pathogens, which simplifies the more complex setting in which multiple respiratory viruses and bacteria circulate simultaneously. We also represented pathogen interaction using a single parameter that modifies susceptibility during infection. In reality, pathogen interactions may be asymmetric, depend on the order and timing of infection, and affect susceptibility, transmissibility, infection duration, or disease severity through different mechanisms. Second, our analysis was based entirely on simulated data, and we have not yet demonstrated the presence or magnitude of hitchhiker bias using empirical surveillance data. The simulations establish that this bias can arise under the specified transmission and observation processes, but they do not show how large it is for any particular pathogen pair or surveillance system. Applying the framework to real-world multiplex testing data will require additional information on community infection prevalence, symptom profiles, healthcare-seeking behavior, testing practices, viral load or infection timing, and the clinical reasons for hospitalization. Third, although we examined a wide range of severity ratios (κ = 1–100), we assumed that each hospitalization was caused by a single pathogen. When two pathogens differ less in severity, both may contribute to the same clinical event, and attributing a hospitalization to one of them may need to be probabilistic rather than absolute. Finally, although we used hospitalizations to illustrate hitchhiker bias, the same mechanism may apply to other forms of symptom-triggered surveillance, including outpatient consultations, emergency-department attendance, and mortality surveillance. Future studies should therefore validate the framework using empirical data and assess its robustness across different pathogens, surveillance systems, and observation processes.

In conclusion, a pathogen detected in a hospitalized patient is not necessarily the cause of the hospitalization. When this distinction is ignored, a mildly pathogenic virus that co-circulates with a more pathogenic one can appear severe, its epidemic can appear to peak earlier, and its interaction with the other virus can appear stronger, absent or even reversed. Mechanistic modeling alone does not remove these distortions; it requires an observation model that separates pathogen-attributable disease from incidental detection. As multiplex testing becomes routine, recognizing and correcting for hitchhiker bias will be important both for reliable inference about pathogen severity and interaction and for public health decisions that depend on such estimates.

## Model and Methods

“Hitchhiker” pathogens are microorganisms present in a diagnostic sample but not responsible for the symptoms that triggered testing. For example, consider a virus (virus 2) that causes few or no symptoms. In symptom-based surveillance, virus 2 would only be detected when an individual co-infected with a different, symptom-causing virus (virus 1) undergoes testing. Consequently, detection of virus 2 becomes conditional on virus 1, systematically mischaracterizing virus 2’s severity.

To formalize the structure of hitchhiker bias, we use the diagram in Fig. 1 to represent how infections, hospitalizations, and observed hospitalizations are linked. In this framework, virus 1 and virus 2 can infect individuals separately or co-infect the same host, and hospital-based multiplex PCR testing is primarily triggered by symptoms caused by virus 1. Asymptomatic infections with virus 2 alone are therefore missed, and virus 2 is observed mainly when it co-occurs with virus 1 in hospitalized patients, creating a biased detection pathway in which the apparent presence of virus 2 reflects its co-occurrence with symptomatic infections rather than its true circulation in the community. The blue arrows in Fig. 1 indicate contributions to observed hospitalizations arising from single infections with each virus, whereas the red arrows indicate additional contributions from co-infected individuals, highlighting how virus 2 “hitchhikes” into the data via hospitalizations driven by virus 1 and vice versa.

### Transmission Model

To investigate hitchhiker bias in hospital-based multiplex surveillance, we used a deterministic compartmental model of two respiratory viruses spreading in a closed population. The model extends the classic SEIR (susceptible–exposed–infectious–recovered) framework to include co-infection: individuals can be infected with either virus alone or with both at the same time [20,42,43]. Both viruses were given identical transmission parameters (basic reproduction number, latent period and infectious period; Table 1), loosely based on SARS-CoV-2. They differed only in their hospitalization probabilities, so any difference between their observed dynamics reflects hitchhiker bias rather than differences in transmissibility.

The state space comprises 16 compartments, each defined by an individual’s joint SEIR status for virus 1 (first letter; rows in Fig. 2) and virus 2 (second letter; columns in Fig. 2). For example, SS denotes individuals susceptible to both viruses, IS individuals infectious with virus 1 and susceptible to virus 2, SI individuals susceptible to virus 1 and infectious with virus 2, and II individuals infectious with both viruses. The full system of ordinary differential equations is given in S1 Text.

Each virus progresses through the SEIR states at its own rates. Susceptible individuals acquire virus i at the force of infection λ_i_, exposed individuals become infectious at rate σ_i_ (mean latent period 1/σ_i_), and infectious individuals recover at rate γ_i_ (mean infectious period 1/γ_i_). The two viruses are coupled through a single interaction parameter, θ, which scales susceptibility to one virus in individuals currently infectious with the other. These individuals acquire virus 2 at rate θλ_2_ and virus 1 at rate θλ_1_ (dotted arrows in Fig. 2). Values of θ > 1 represent facilitation, θ < 1 represents inhibition, and θ = 1 means no interaction. This formulation captures biologically plausible within-host effects during an ongoing infection: innate immune responses may reduce susceptibility to a second virus, whereas damage to the respiratory epithelium may increase it. We first simulated epidemics without interaction (θ = 1) to test whether, in the presence of hitchhiker bias, standard model-fitting approaches infer a spurious interaction (θ *≠* 1). We then simulated an inhibitory (θ = 0.5) and a facilitative (θ = 2) interaction to test whether a true interaction can still be recovered.

### Symptom-based Surveillance

To simulate surveillance, we assume that only infectious individuals who require hospitalization are eligible for testing. Each virus has an associated probability of hospitalization (*ρ*_1_ for virus 1 and *ρ*_*2*_ for virus 2), representing the probability that an infected individual will be hospitalized due to infection and thus captured in surveillance data. Testing is applied only to compartments containing infectious individuals (i.e., SI, IS, EI, IE, II, IR and RI). All hospitalized patients are tested for both viruses with a multiplex panel of perfect sensitivity and specificity, and individuals who are exposed but not yet infectious test negative. Infections that do not result in hospitalization remain undetected unless accompanied by co-infection with a virus that does lead to hospitalization. This approach mimics real-world hospital-based surveillance systems, where case detection is strongly biased toward individuals with more severe illness.

### True and observed Hospitalizations

Equations for the true hospitalizations from virus 1 and virus 2 are:

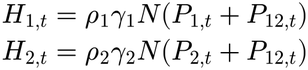

Where *ρ*_*1*_ and *ρ*_*2*_ is the proportion of infection resulting in hospitalizations.

Here,*P*_*1,t*_, *P*_*2,t*_, and *P*_*12,t*_ are the prevalences of mono-infection and co-infection with virus 1 and virus 2, given by:

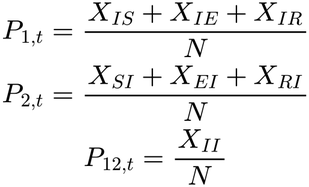

Here, *X*_*ij*_ represents the number of individuals in state *i* for virus 1 and *j* for virus 2, and is the total population size. The equations for the remaining compartments are in the S1 Text.

Because every hospitalized patient is tested for both viruses, the number of patients who test positive for a virus can exceed the number hospitalized because of it. We define the observed hospitalizations, 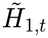 and 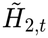, as the numbers of hospitalized patients who test positive for virus 1 and virus 2, respectively (Fig. 1):

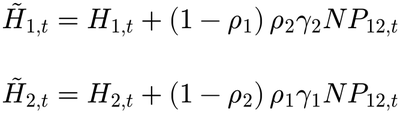

The first term of each equation is the true hospitalizations caused by that virus. The second term is the hitchhiker component: co-infected patients hospitalized because of the other virus, in whom the focal virus is detected incidentally. For virus 2, for example *ρ*_1_*γ*_1_*NP*_12,*t*_, is the number of co-infected patients hospitalized because of virus 1. The factor (1 − ρ_2_) excludes those who would also have been hospitalized because of virus 2, since they are already counted in *H*_2,*t*_. We therefore assume that each hospitalization is attributed to a single cause.

In this study, hospitalizations arise only from infection with the two modeled viruses. In practice, patients are also hospitalized because of other respiratory and non-respiratory pathogens, as well as for non-infectious reasons. Any such patient who is tested with a multiplex panel could provide an additional opportunity for incidental detection of virus 1 or virus 2. Because these background hospitalizations were not modeled, our simulations capture only the hitchhiker pathway generated by co-infection between the two modeled viruses. In settings where other pathogens cause a substantial share of hospitalizations, the number of incidental detections, and therefore the magnitude of hitchhiker bias, may be larger than estimated here.

To evaluate the presence and impact of hitchhiker bias, we simulated the spread of two viruses that are identical in all epidemiological parameters except for their initial number of exposed individuals and hospitalization probabilities. Specifically, we varied the initial number of exposed individuals for virus 1 from *N*/10^12^ from to *N*/10^2^, while keeping that of virus 2 fixed at *N*/10^12^. This setup produced differing degrees of temporal overlap between the infection curves of virus 1 (*I*_*1*_) and virus 2 (*I*_*2*_), ranging from minimally overlapping to fully overlapping epidemics. Here and are defined as

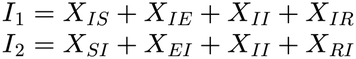

To isolate the influence of hitchhiker bias, we first assigned contrasting hospitalization probabilities: virus 1 was designated as more pathogenic with a high hospitalization probability (*ρ*_1_ = 0.05), while virus 2 was largely asymptomatic or mild, with a low hospitalization probability (*ρ*_2_ = 0.0005). All other transmission and recovery parameters were held constant. This design allows us to investigate how virus 2, acting as a potential hitchhiker pathogen, may be disproportionately detected due to co-infection with the more severe virus 1.

### Simulation details

To simulate the deterministic trajectory of the model, we used the pomp package (version 5.11) [44]. The pomp model object was initialized with biologically plausible parameter values chosen to represent two co-circulating respiratory viruses. Specifically, the basic reproduction numbers for both viruses were set to 2.5, with average latent and infectious periods of 4 and 5 days, respectively. Initial conditions included no recovered individuals, initially *N*/10^12^ exposed individuals for virus 2, and between *N*/10^12^ and *N*/10^2^ initially exposed individuals for virus 1. The hospitalization probability was set to 0.05 for virus 1 and 0.0005 for virus 2, reflecting their differing pathogenicity profiles. These parameters are also listed in Table 1.

## Model fitting

### Observation model

To account for variability and uncertainty in the number of individuals hospitalized, we modeled the observation process using a negative binomial distribution, which naturally accommodates the overdispersion often observed in epidemiological surveillance data. At each time point, the observed number of hospitalizations was assumed to follow a negative binomial distribution with mean equal to the model-predicted number of new hospitalizations and a dispersion parameter *ψ*. Specifically, if *y*_*i,t*_, denotes the observed number of hospitalizations positive for virus *i* on day *t*, then

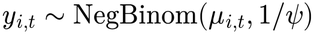

where *μ*_*i*_,_*t*_ is the expected number of new hospitalizations and 1/*ψ* is the size parameter used in the implementation. Under this parameterization,

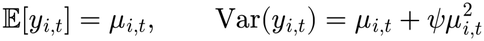

Thus, *ψ* directly controls the level of overdispersion: as *ψ* → 0, the variance approaches the mean (recovering the Poisson case), while larger values of *ψ* allow the variance to grow quadratically with the mean.

To reflect context-specific contributions to observed cases, we incorporated a covariate representing the hitchhiker effect. When the hitchhiker effect is modeled (*h*_*t*_ = 1), the expected observations are the observed hospitalizations 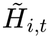; otherwise, they are the true hospitalizations *H*_*i,t*_.

The mean *μ*_*i*_,_*t*_ is determined by the hitchhiker effect covariate *h*_*t*_, such that:

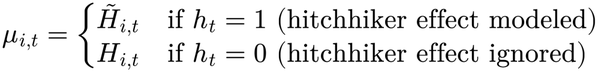

For this simulation study, we focused on estimating six main parameters of the model: the basic reproduction numbers of virus 1 and virus 2 (ℛ_*i*1_, ℛ_*i*2_), their hospitalization probabilities (*ρ*_1_, *ρ*_2_), the interaction strength between the two viruses (θ) and the overdispersion parameter *ψ*.

### MCMC

Model fitting was accomplished using a Bayesian MCMC approach implemented in the BayesianTools package in R. More specifically, we employed differential evolution with snooker updates (DEzs) as the sampling algorithm, which has been shown to perform well for correlated posterior distributions with few chains [45,46]. Prior distributions and fitting settings are listed in S1 Table. Starting values for all chains were drawn at random from the prior distributions, so that chains started from dispersed points in parameter space, as required for multi-chain convergence diagnostics [47]. Five independent chains were then run for 20,000 iterations each, with the first half of samples discarded as burn-in to reduce the influence of initialization. Posterior samples from the remaining iterations were pooled to summarize marginal distributions and estimate posterior means and 95% credible intervals for each parameter. We computed the Gelman–Rubin potential scale reduction factor (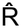) for each parameter and excluded replicates with 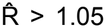 for any parameter. We visually inspected trace plots for evidence of mixing across chains and examined pairwise posterior correlation plots to evaluate identifiability and dependencies between parameters.

### Numerical implementation

All model fitting and diagnostics were carried out in R (version 4.4.2). Likelihoods were specified using the pomp package (version 5.11), sampling was implemented with BayesianTools (version 0.1.8), and posterior summaries and diagnostic plots were generated with MCMCvis [48].

## Data Availability

The code used in this study is currently available on a private GitHub repository and will be made publicly available upon publication.

## Acknowledgments

We thank members of the Infectious Disease Epidemiology lab for a group discussion of this study. The study was funded by the Max Planck Society.

