## Supplementary materials for "Hitchhiker bias can distort inference of pathogen severity and pathogen–pathogen interactions from symptom-based surveillance: A modeling study"

### S1 Text. Model equations.

The dynamics of virus 1 and virus 2 are described by a system of ordinary differential equations (ODEs) for the number of individuals $X_{ij}(t)$ in state $i$ for virus 1 and state $j$ for virus 2, where $i, j \in \{S, E, I, R\}$ (susceptible, exposed, infectious, recovered) and $t$ is time in days. The total population size $N$ is constant.

Equations for the 1st column in the model schematic:

$$\begin{aligned}
\frac{dX_{SS}}{dt} &= -(\lambda_1 + \lambda_2)\, X_{SS} \\
\frac{dX_{ES}}{dt} &= \lambda_1 X_{SS} - (\sigma_1 + \lambda_2)\, X_{ES} \\
\frac{dX_{IS}}{dt} &= \sigma_1 X_{ES} - (\gamma_1 + \theta\lambda_2)\, X_{IS} \\
\frac{dX_{RS}}{dt} &= \gamma_1 X_{IS} - \lambda_2 X_{RS}
\end{aligned}$$

Equations for the 2nd column in the model schematic:

$$\begin{aligned}
\frac{dX_{SE}}{dt} &= \lambda_2 X_{SS} - (\lambda_1 + \sigma_2)\, X_{SE} \\
\frac{dX_{EE}}{dt} &= \lambda_2 X_{ES} + \lambda_1 X_{SE} - (\sigma_1 + \sigma_2)\, X_{EE} \\
\frac{dX_{IE}}{dt} &= \theta\lambda_2 X_{IS} + \sigma_1 X_{EE} - (\gamma_1 + \sigma_2)\, X_{IE} \\
\frac{dX_{RE}}{dt} &= \lambda_2 X_{RS} + \gamma_1 X_{IE} - \sigma_2 X_{RE}
\end{aligned}$$

Equations for the 3rd column in the model schematic:

$$\begin{aligned}
\frac{dX_{SI}}{dt} &= \sigma_2 X_{SE} - (\theta\lambda_1 + \gamma_2)\, X_{SI} \\
\frac{dX_{EI}}{dt} &= \sigma_2 X_{EE} + \theta\lambda_1 X_{SI} - (\sigma_1 + \gamma_2)\, X_{EI} \\
\frac{dX_{II}}{dt} &= \sigma_2 X_{IE} + \sigma_1 X_{EI} - (\gamma_1 + \gamma_2)\, X_{II} \\
\frac{dX_{RI}}{dt} &= \sigma_2 X_{RE} + \gamma_1 X_{II} - \gamma_2 X_{RI}
\end{aligned}$$

Equations for the 4th column in the model schematic:

$$\begin{aligned}
\frac{dX_{SR}}{dt} &= \gamma_2 X_{SI} - \lambda_1 X_{SR} \\
\frac{dX_{ER}}{dt} &= \gamma_2 X_{EI} + \lambda_1 X_{SR} - \sigma_1 X_{ER}
\end{aligned}$$

$$\frac{dX_{IR}}{dt} = \gamma_2 X_{II} + \sigma_1 X_{ER} - \gamma_1 X_{IR}$$
$$\frac{dX_{RR}}{dt} = \gamma_2 X_{RI} + \gamma_1 X_{IR}$$

Here, $\lambda_1(t)$ and $\lambda_2(t)$ are the forces of infection of virus 1 and virus 2, respectively: $\lambda_1 = \mathcal{R}_{n1}\,\gamma_1\left(P_1(t) + P_{12}(t)\right)$ $\lambda_2 = \mathcal{R}_{n2}\,\gamma_2\left(P_2(t) + P_{12}(t)\right)$ where $\sigma_j$ and $\gamma_j$ are the rates of progression to the infectious state and of recovery for virus $j$, and $\theta$ scales susceptibility to one virus while infectious with the other. $R_{n1}$ and $R_{n2}$ are the effective reproduction numbers defined as:

$$\mathcal{R}_{n1} = \frac{\mathcal{R}_{i1}}{1 - R_{10}}, \quad \mathcal{R}_{n2} = \frac{\mathcal{R}_{i2}}{1 - R_{20}}$$

Here, $P_{1,t}$, $P_{2,t}$ and $P_{12,t}$ are the prevalences of mono-infection and co-infection with virus 1 and virus 2, given by:

$$P_{1,t} = \frac{X_{IS} + X_{IE} + X_{IR}}{N}$$

$$P_{2,t} = \frac{X_{SI} + X_{EI} + X_{RI}}{N}$$

$$P_{12,t} = \frac{X_{II}}{N}$$

Here, $X_{ij}$ represents the number of individuals in the state $i$ for virus 1 and $j$ for virus 2, and $N$ is the total population size.

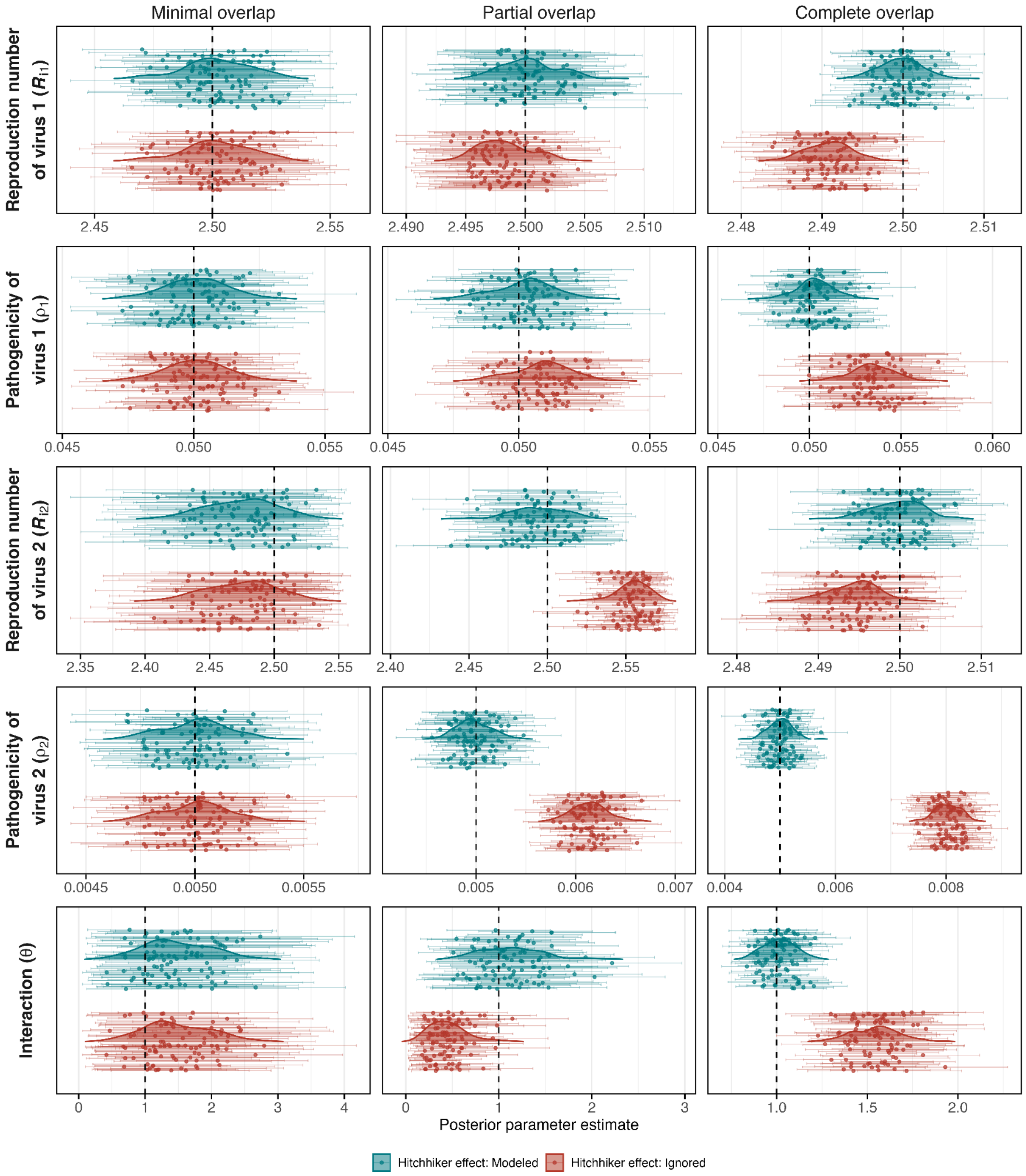


**S1 Fig. Parameter estimates in the absence of interaction for $\rho_2$ = 0.005 ($\kappa$ = 10).** Posterior estimates obtained by fitting models to simulated epidemics with minimal, partial, and complete epidemic overlap (columns), generated without interaction ($\theta$ = 1) and with $\rho_1$ = 0.05. Rows show the basic reproduction number of virus 1, the pathogenicity of virus 1, the basic reproduction number of virus 2, the pathogenicity of virus 2, and the interaction parameter. Models in which the hitchhiker effect was modeled (teal) or ignored (red) were fitted to 100 simulated datasets per scenario. Each point is the posterior mean for one simulated dataset, and horizontal bars show the corresponding 95% credible interval. Shaded density curves summarize the distribution of posterior means across datasets. Vertical dashed lines indicate the true parameter values.

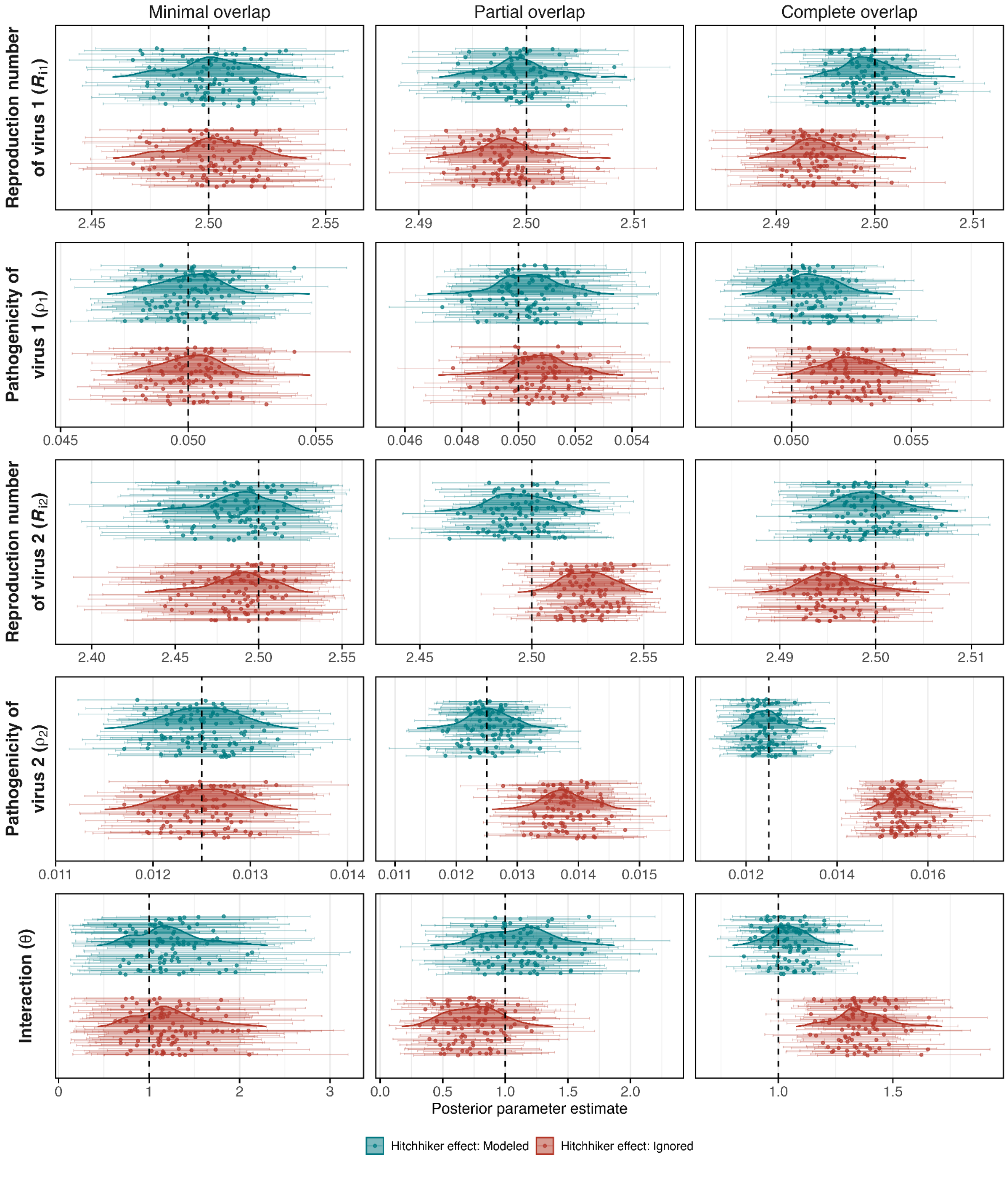


**S2 Fig. Parameter estimates in the absence of interaction for $\rho_2$ = 0.0125 ($\kappa$ = 4).** Posterior estimates obtained by fitting models to simulated epidemics with minimal, partial, and complete epidemic overlap (columns), generated without interaction ($\theta$ = 1) and with $\rho_1$ = 0.05. Rows show the basic reproduction number of virus 1, the pathogenicity of virus 1, the basic reproduction number of virus 2, the pathogenicity of virus 2, and the interaction parameter. Models in which the hitchhiker effect was modeled (teal) or ignored (red) were fitted to 100 simulated datasets per scenario. Each point is the posterior mean for one simulated dataset, and horizontal bars show the corresponding 95% credible interval. Shaded density curves summarize the distribution of posterior means across datasets. Vertical dashed lines indicate the true parameter values.

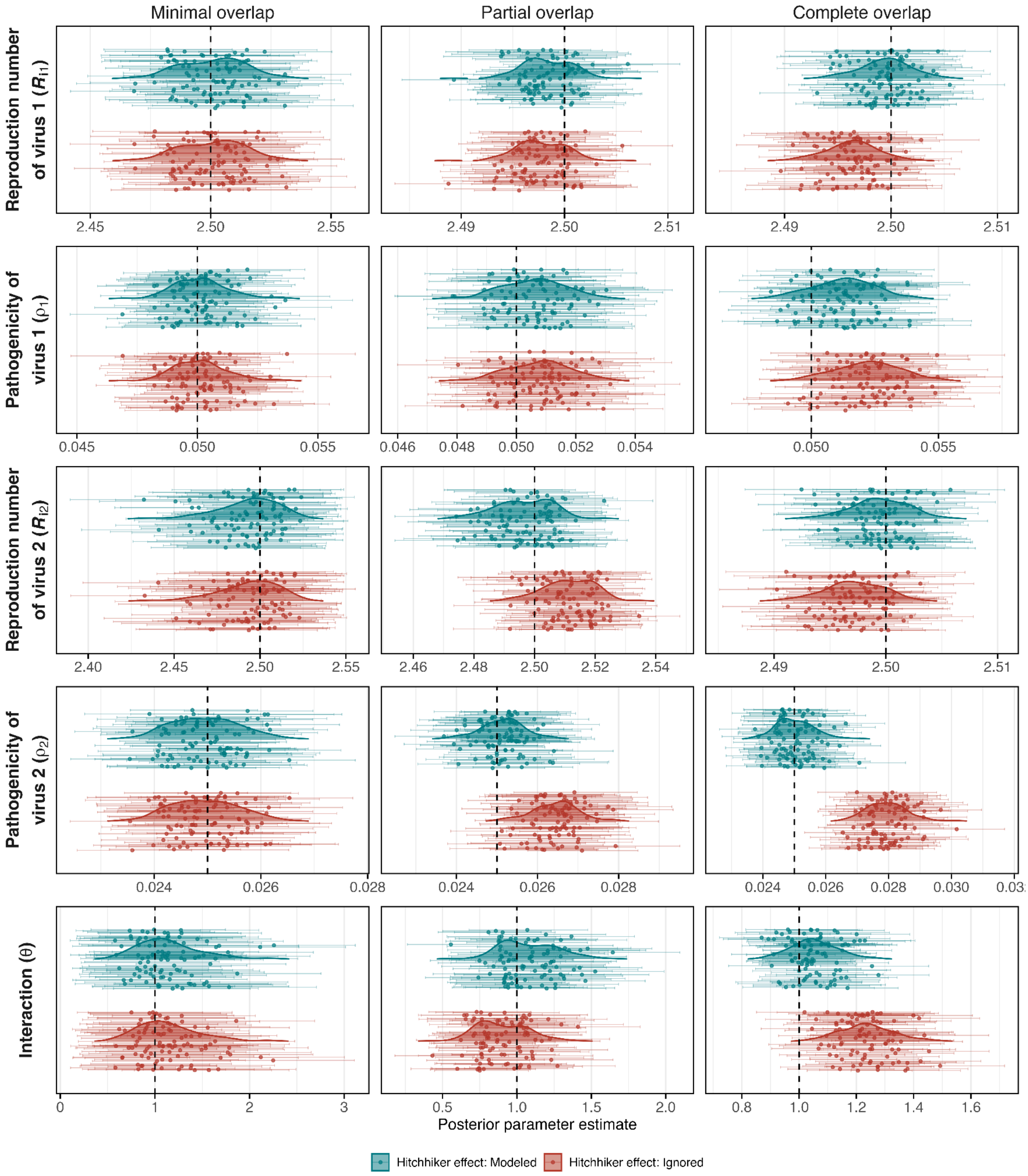


**S3 Fig. Parameter estimates in the absence of interaction for $\rho_2$ = 0.025 (κ = 2).** Posterior estimates obtained by fitting models to simulated epidemics with minimal, partial, and complete epidemic overlap (columns), generated without interaction (θ = 1) and with $\rho_1$ = 0.05. Rows show the basic reproduction number of virus 1, the pathogenicity of virus 1, the basic reproduction number of virus 2, the pathogenicity of virus 2, and the interaction parameter. Models in which the hitchhiker effect was modeled (teal) or ignored (red) were fitted to 100 simulated datasets per scenario. Each point is the posterior mean for one simulated dataset, and horizontal bars show the corresponding 95% credible interval. Shaded density curves summarize the distribution of posterior means across datasets. Vertical dashed lines indicate the true parameter values.

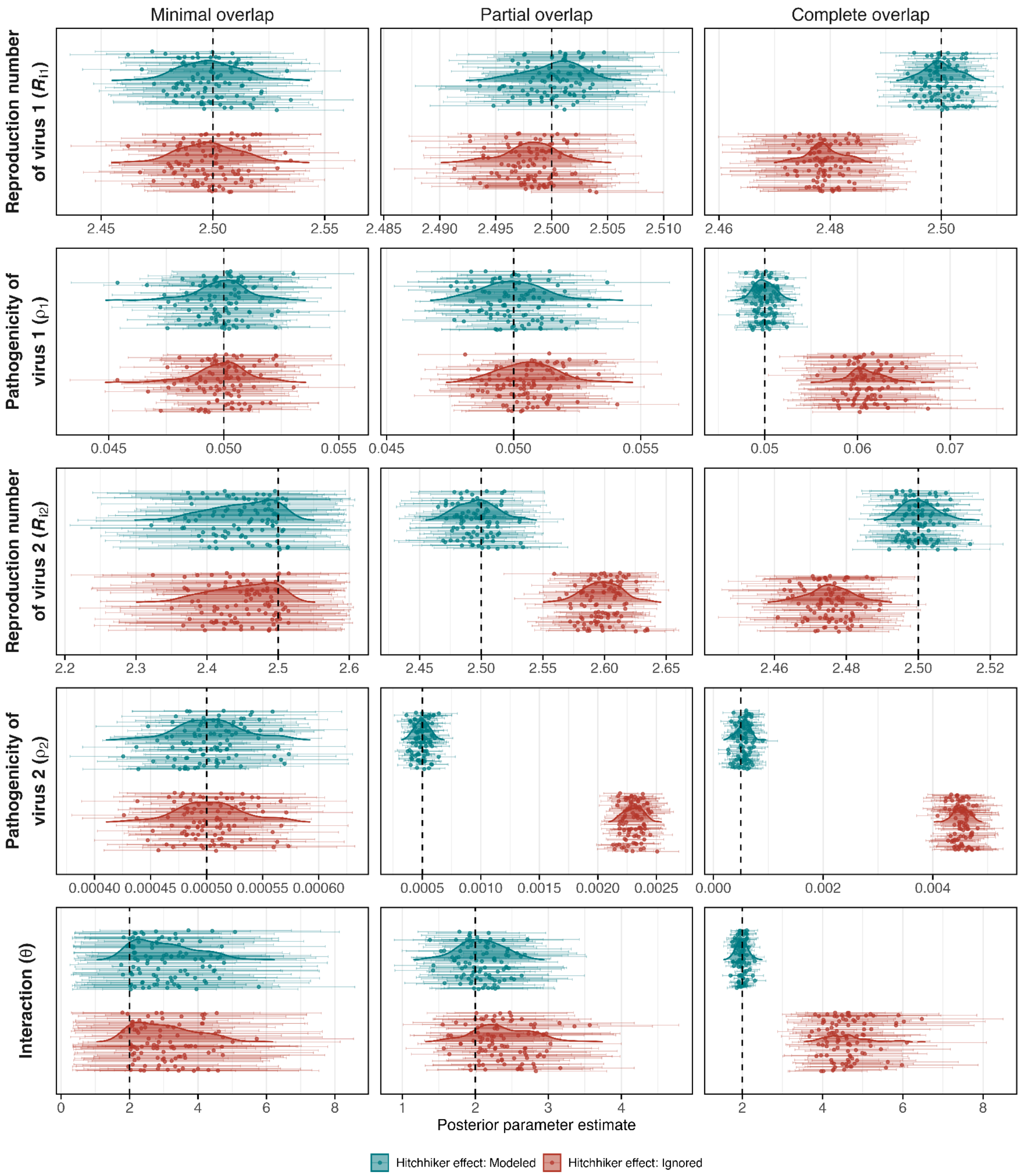


**S4 Fig. Parameter estimates under a positive interaction ($\theta$ = 2).** Posterior estimates obtained by fitting models to simulated epidemics with minimal, partial, and complete epidemic overlap (columns), generated with a positive interaction ($\theta$ = 2), $\rho_1$ = 0.05 and $\rho_2$ = 0.0005. Rows show the basic reproduction number of virus 1, the pathogenicity of virus 1, the basic reproduction number of virus 2, the pathogenicity of virus 2, and the interaction parameter. Models in which the hitchhiker effect was modeled (teal) or ignored (red) were fitted to 100 simulated datasets per scenario. Replicates whose chains did not converge ($\hat{R}$ > 1.05 for any parameter) were excluded. Each point is the posterior mean for one simulated dataset, and horizontal bars show the corresponding 95% credible interval. Shaded density curves summarize the distribution of posterior means across datasets. Vertical dashed lines indicate the true parameter values.

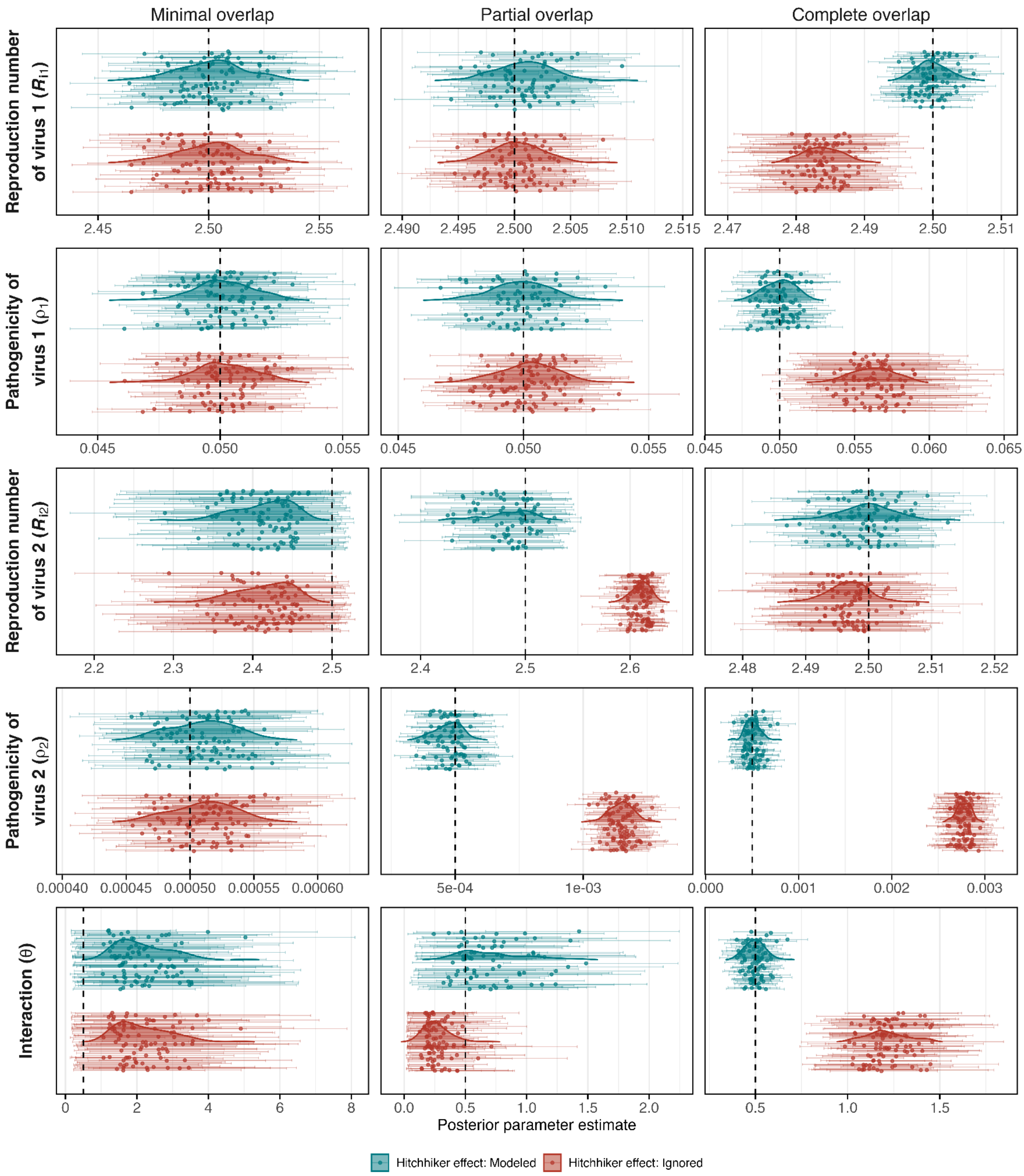


**S5 Fig. Parameter estimates under a negative interaction (θ = 0.5).** Posterior estimates obtained by fitting models to simulated epidemics with minimal, partial, and complete epidemic overlap (columns), generated with a negative interaction (θ = 0.5), $\rho_1$ = 0.05 and $\rho_2$ = 0.0005. Rows show the basic reproduction number of virus 1, the pathogenicity of virus 1, the basic reproduction number of virus 2, the pathogenicity of virus 2, and the interaction parameter. Models in which the hitchhiker effect was modeled (teal) or ignored (red) were fitted to 100 simulated datasets per scenario. Replicates whose chains did not converge ($\hat{R}$ > 1.05 for any parameter) were excluded. Each point is the posterior mean for one simulated dataset, and horizontal bars show the corresponding 95% credible interval. Shaded density curves summarize the distribution of posterior means across datasets. Vertical dashed lines indicate the true parameter values.

**S1 Table. Specification of the Bayesian inference procedure.**

| Component | Parameter | Specification |
|---|---|---|
| **Data** | Fitted data | Daily observed hospitalizations of virus 1 and virus 2, $\tilde{H}_1(t)$ and $\tilde{H}_2(t)$, fitted jointly |
| | Simulated data | 100 datasets per scenario, generated with negative binomial noise ($\psi$ = 0.04) |
| **Likelihood** | $y_{i,t}$ | Negative binomial with mean $\mu_{i,t}$ and size $1/\psi$, so $Var(y_{i,t}) = \mu_{i,t} + \psi\mu_{i,t}^2$; log-likelihoods summed over both viruses and all days |
| **Priors (independent)** | $R_{01}$, $R_{02}$ | $R_{0i}/10$ ~ Exponential(0.1), truncated to [0.1, 1], i.e. $R_{0i} \in [1, 10]$ |
| | $\rho_1$, $\rho_2$ | Beta(2, 2) on [0, 1] |
| | $\theta$ | $\theta/10$ ~ Beta(2, 2), i.e. $\theta \in [0, 10]$ |
| | $\psi$ | Uniform(0, 1) |
| **Fixed parameters** | Latent period | $\sigma_1 = \sigma_2 = 1/4$ day$^{-1}$ |
| | Infectious period | $\gamma_1 = \gamma_2 = 1/5$ day$^{-1}$ |
| | Population size | $N = 10^6$ |
| | Initial exposed | $E_1(0) = 10^4$, $10^{-4}$ or $10^{-6}$ (minimal, partial and complete overlap); $E_2(0) = 10^{-6}$ |
| | Other | No initial immunity; no seasonal forcing |
| **Computation** | Model solution | Deterministic ODE model implemented in pomp and solved with deSolve |
| | Sampler | Differential evolution MCMC with snooker updates (DEzs), BayesianTools |
| | Chains | 5 chains × 20,000 iterations; first 10,000 discarded as burn-in |
| | Convergence | Replicates with $\hat{R} > 1.05$ excluded |
